# Whole Exome Sequencing in dense families suggests genetic pleiotropy amongst Mendelian and complex neuropsychiatric syndromes

**DOI:** 10.1101/2021.11.04.21265926

**Authors:** Suhas Ganesh, Alekhya Vemula, Samsiddhi Bhattacharjee, Kezia Mathew, Dhruva Ithal, Karthick Navin, Ravi Kumar Nadella, Biju Viswanath, Patrick F Sullivan, The ADBS consortium, Sanjeev Jain, Meera Purushottam

**Affiliations:** Central Institute of Psychiatry, Kanke, Ranchi, India; Schizophrenia Neuropharmacology Research Group, Department of Psychiatry, Yale University School of Medicine, New Haven, US; Department of Psychiatry, National Institute of Mental Health and Neuro Sciences, Bengaluru, India; National Institute of Biomedical Genomics, Kalyani, India; Department of Psychiatry, Varma Hospital, Bhimavaram; India; FRANZCP, Psychiatry, University of North Carolina at Chapel Hill, United States; Department of Medical Epidemiology and Biostatistics at Karolinska Institutet, Sweden; Department of Psychiatry, National Institute of Mental Health and Neuro Sciences, Bengaluru

## Abstract

Whole Exome Sequencing (WES) studies provide important insights into the genetic architecture of serious mental illness (SMI). Genes that are central to the shared biology of SMIs may be identified by WES in families with multiple affected individuals with diverse SMI (F-SMI). We performed WES in 220 individuals from 75 F-SMI families and 60 unrelated controls. Within pedigree prioritization employed criteria of *rarity, functional consequence*, and *sharing* by ≥3 affected members. Across the sample, gene and gene-set-wide case-control association analysis was performed with Sequence Kernel Association Test, accounting for kinship.

In 14/16 families with ≥3 affected individuals, we identified a total of 79 rare predicted deleterious variants in 79 unique genes shared by ≥3 members with SMI and absent in 60 unrelated controls. Twenty (25%) genes were implicated in monogenic neurodevelopmental syndromes in OMIM, a fraction that is a significant overrepresentation (Fisher’s Exact test OR = 2.47, p = 0.001). In gene-set wise SKAT, statistically significant association was noted for genes related to synaptic function (SKAT-p = 0.014). In this WES study in F-SMI, we identify private, rare, protein altering variants in genes previously implicated in Mendelian neuropsychiatric syndromes; suggesting pleiotropic influences in neurodevelopment between complex and Mendelian syndromes.

## Introduction

Neuropsychiatric syndromes, such as schizophrenia (SCZ), bipolar disorder (BD), obsessive compulsive disorder (OCD) and substance use disorders (SUDs) (referred hereafter as serious mental illness [SMI]) often cluster in families ^1,2^. Next generation sequencing (NGS) can be used to explore the genetics of complex, common disorders, using both case-control and family-based designs ^3^. These include common variant genome wide association studies (GWAS) and exome wide studies that scan for rare coding variants ^4,5^. Such methods have provided useful details about the contribution of rare variants to the genetic architecture of SMI, often complementing the common variant contributions identified in GWAS.

Case-control studies typically exclude relatives to minimize sampling bias and potential false positive associations. About 10% of persons affected with SMI have an affected first degree relative. The increased occurrence of potentially disease relevant variants, in densely affected pedigrees, may offer clues towards genes and pathways involved in the neurobiology of SMI. Many studies employing whole exome sequencing (WES) in psychiatry have analyzed families with multiple affected members (reviewed in ^6^). These studies have generally focused on pedigrees with a cluster of individuals affected with a specific SMI such as BD or SCZ. Family-based studies in SCZ ^7-9^, BD ^10-13^ and OCD ^14^ have identified multiple rare *de novo* and loss of function variations relevant to the biology of each syndrome. Similarly, case-control association studies in SCZ ^4,5^ and BD ^15,16^ have also identified the contribution of rare variants of large effect, advancing the current understanding of genetic architecture of these syndromes. An overview of recent findings from family based and case-control sequencing studies in SMI is presented in the supplement S1.

In summary, we detect a trend towards a higher burden of rare, protein altering variations, across cases with different SMI syndromes, when compared to controls ^17,18^, although some studies are equivocal ^12,15^. These variants tend to be overrepresented in genes that are integral to neurodevelopment and synaptic function, and these ontologies are often identified across SMI syndromes ^19-21^. Often, some genes with a higher burden of rare variants, identified in family studies, have also been implicated in other severe neuropsychiatric syndromes, with features of anomalies in neurodevelopment and neurodegeneration ^22,23^. There seems to be considerable overlap of rare variants that contribute to the risks of SMI, across syndromes. This convergence has also been observed for common variants ^24^, as well as across genes, gene networks ^25,26^, molecular pathways ^27^, and brain imaging endophenotypes ^28^. A recent analyses that integrated GWAS findings across 11 major psychiatric syndromes, suggests a shared genetic architecture across syndromes at bio-behavioural, functional genomic and molecular genetic levels ^29^. A co-aggregation of SMI syndromes, and overlapping symptom dimensions, are often seen within a family ^1,2^. Rare variants, identified in families with multiple ill members, may thus may explain a proportion of the risk in the population. They are obviously of great heuristic value, to explore the pathobiology of the disease, and correlates of clinical features.

In this study, we examined the occurrence of rare, deleterious variants in individuals from families that had multiple affected members (as identified in Accelerator Program for Discovery in Brain Disorders using Stem Cells (ADBS)) ^30^. Within pedigree segregation, as well as cross-sample case-control association tests, were used to prioritize risk variants. We examined the functional and clinical significance of the prioritized genes and variants, including evolutionary conservation, mutation intolerance, brain expression, protein function and disease relevance. Specifically, we examined if the genes carrying the prioritized variants are overrepresented in Mendelian neuropsychiatric syndromes, and synaptic genes (as attempted for SCZ in the Xhosa population ^4^). In families segregating a variant in a gene linked to a syndrome, we also reviewed the clinical profile of affected individuals for symptoms and signs of the particular Mendelian syndrome.

## Results

### Sample

We sequenced 280 (131 females) individuals, including 220 from 75 families (F-SMI), and 60 unrelated unrelated-controls. Of the F-SMI samples, 160 were cases diagnosed with a SMI: SCZ (n = 63), BD (n = 80), OCD (n = 7), SUD (n = 7), complex SMI (n = 3) along with 60 family-controls without a lifetime diagnosis of mental illness. The demographic profile of the sample with age and sex distribution and illness profile of the affected sample is provided (Table 1). Fifty-one (68%) of the 75 families had cases with diagnosis across ≥2 of the 4 categories noted above.

**Table 1:** Demographic and profile of the sample.

| **1a. Demographic profile of cases, family controls and unrelated controls** |  |  |  |  |  |
| --- | --- | --- | --- | --- | --- |
| **Groups** | **Cases** | **Family Controls** | **Unrelated Controls** | **F/Chi** | **p** |
| Sample size | 160 | 60 | 60 |  |  |
| Age - mean (SD) | 39(13.5) | 43.8(15.3) | 45.6(18.2) | 5.13 | 0.006 |
| Sex male/female | 92/68 | 28/32 | 29/31 | 2.79 | 0.25 |
| **1b. Clinical profile by diagnosis** |  |  |  |  |  |
| **Diagnosis** | **Sample size** | **Age mean (SD)** | **Sex (male/female)** | **AAO mean (SD)** | **DOI mean (SD)** |
| Schizophrenia | 63 | 37.1(12.9) | 35/28 | 23.1(7.5) | 12.4(9.8) |
| Bipolar disorder | 80 | 39.9(13.5) | 45/35 | 21.9(7.5) | 19.9(11.3) |
| OCD | 7 | 33.7(12.1) | 2/5 | 24.4(7.7) | 8.4(7.3) |
| Addiction | 7 | 42.9(13.9) | 7/0 | 33.8(14.4) | 9(7.9) |
| Mixed SMI | 3 | 46 (22.9) | 3/0 | 41.6 (14.2) | 8(8.2) |
| AAO – Age at onset, DOI – Duration of illness, SD – standard deviation |  |  |  |  |  |

Within the F-SMI familial sample, the median number of samples from cases per family were 2 (range:1 – 6) and family-controls was 1 (range:1 – 7). A subset of 16 multi-sample F-SMI (≥3 exome samples) was selected for analysis of within pedigree segregation of putatively deleterious variants.

### Variant profile

Variant calling the exomes of 280 samples resulted in identification of 793818 unique variants. Among these, the median (IQR) number of synonymous and non-synonymous SNVs per sample were 9279 (2146) and 8721 (2433) respectively. The median (IQR) frequency of rare variants (MAF < 0.1%) per sample was 717 (515). A breakdown of the variant profile in the sample is provided in Figure 1.

**Figure 1:**
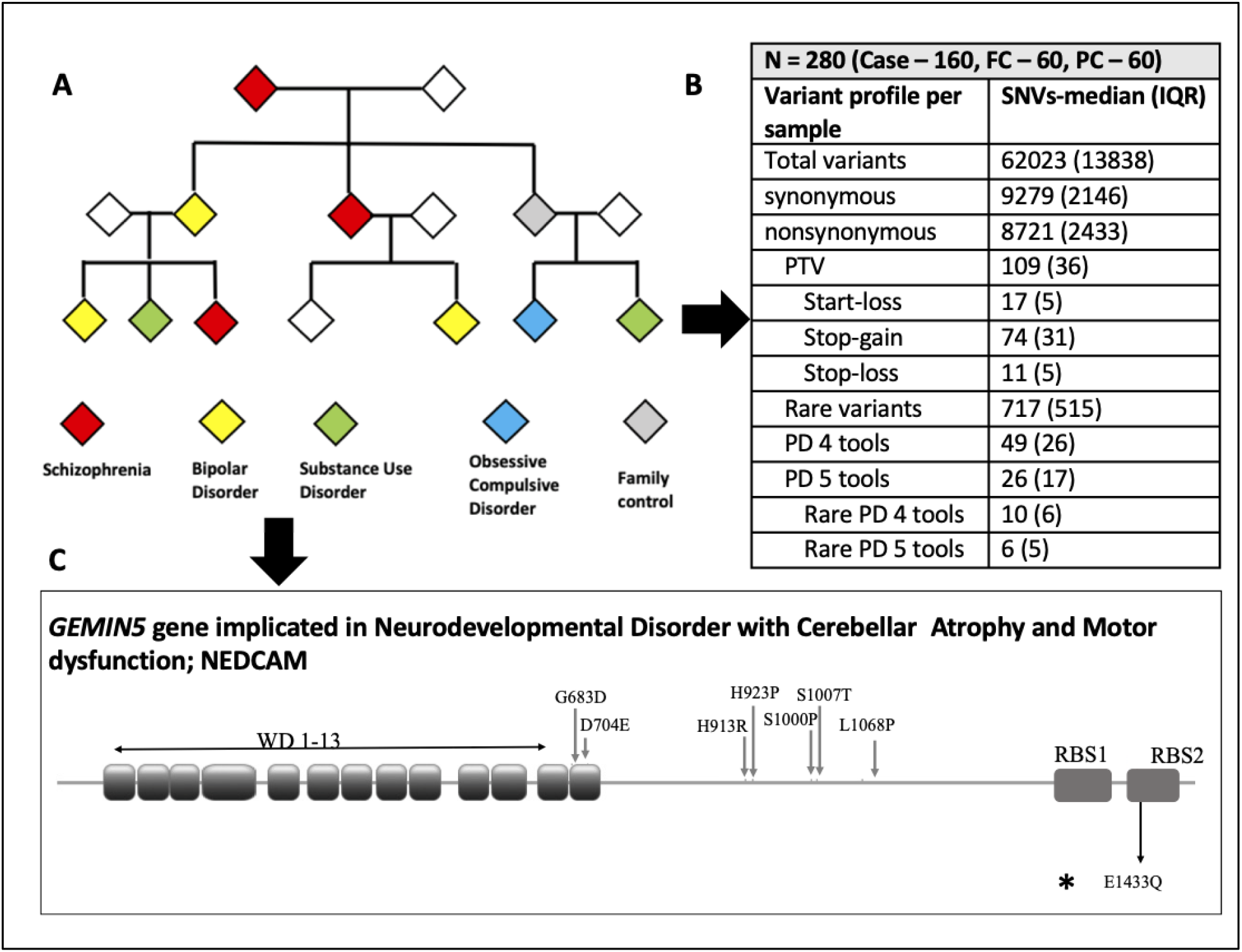
Conceptual overview **-** Exome sequencing was done for individuals from families with multiple members with serious mental illness (SMI). The Table shows the meaningful (significant) variants detected. Details of one representative gene (GEMIN5) are shown. The seven variants marked above have been implicated in Neurodevelopmental Disorder with Cerebellar Atrophy and Motor dysfunction: NEDCAM earlier. The variant marked below **rs544452250** (5-154268943-C-G; E1433Q) was predicted to be deleterious, and was shared across three individuals with schizophrenia, BPAD and Substance use disorder, in one family.

### Within pedigree segregation of private variants

Among 14 of the 16 multi-sample high-density families, we identified a total of 79 RPD variants in 79 different genes that were shared by ≥3 affected members within a family and were absent in the 60 unrelated-controls (Supplementary Table 1). Of these, 78 variants were private to one among the 14 pedigrees. The variant at the position chr5:154268943 in *GEMIN5* gene was shared by 3/6 and 3/3 cases from families D002 and D012 respectively (Figure 1, Table 2). In the remaining set of 59 pedigrees with ≤2 case samples, 15 of the same 79 genes were noted to carry RPD variants that were absent in the 60 unrelated-controls (Supplementary Table 2). Additionally, at the variant level, 4 of the same 79 RPD variants prioritized from the of 14 families, were also noted in this latter set of 59 families (Supplementary Table 3).

**Table 2:** Variants segregating within families with SMI in genes implicated in Mendelian Syndromes.

| chr:position | Reference<br>> alternate | Gene | Family Number | Burden<br>- case | Gene description | OMIM syndrome |
| --- | --- | --- | --- | --- | --- | --- |
| chr18:12337480 | G>C | **AFG3L2** | D002 | 3 | AFG3 like matrix AAA peptidase subunit 2 | Spinocerebellar Ataxia 28; SCA28 |
| chr17:3384920 | A>G | **ASPA** | D002 | 3 | aspartoacylase | Canavan-Van Bogaert-Bertrand disease - spongy degeneration of central nervous system |
| chr9:141016353 | C>T | **CACNA1B** | D002 | 4 | calcium voltage-gated channel subunit alpha1 B | Neurodevelopmental disorder with seizures and nonepileptic hyperkinetic movements |
| chr11:64953762 | G>A | **CAPN1** | D002 | 3 | calpain 1 | Spastic paraplegia 76, autosomal recessive |
| chr20:10625568 | C>G | **JAG1** | D002 | 5 | jagged canonical Notch ligand 1 | Alagille syndrome 1 |
| chr1:10035801 | C>T | **NMNAT1** | D002 | 3 | nicotinamide nucleotide adenyltransferase 1 | Spondyloepiphyseal dysplasia, sensorineural hearing loss, impaired intellectual development, and leber congenital amaurosis; shilca |
| chr5:154268943 | C>G | **GEMIN5** | D002 | 3 | gem nuclear organelle associated protein 5 | Neurodevelopmental disorder with cerebellar atrophy and motor dysfunction; NEDCAM |
| chr5:154268943 | C>G | **GEMIN5** | D012 | 3 | gem nuclear organelle associated protein 5 | Neurodevelopmental disorder with Cerebellar Atrophy and Motor dysfunction; NEDCAM |
| chr21:47860904-47860904 | - > G | **PCNT** | D003 | 6 | pericentrin | Microcephalic osteodysplastic primordial dwarfism, type II |
| chr12:120241184 | G>A | **CIT** | D004 | 3 | citron rho-interacting serine/threonine kinase | microcephaly 17 |
| chr10:70225532 | G>A | **DNA2** | D006 | 4 | DNA replication helicase/nuclease 2 | seckel syndrome 8 |
| chr2:166032822 | G>A | **SCN3A** | D006 | 4 | sodium voltage-gated channel alpha subunit 3 | Epilepsy Familial Focal With Variable Foci 4, Developmental and Epileptic Encephalopathy 62 |
| chr12:23998998 | C>T | **SOX5** | D006 | 4 | SRY-box transcription factor 5 | Lamb Shaffer Syndrome |
| chr2:69409769 | G>T | **ANTXR1** | D007 | 3 | ANTXR cell adhesion molecule 1 | Growth retardation, alopecia, pseudoanodontia, and optic atrophy |
| chr17:40715328 | C>T | **COASY** | D007 | 3 | Coenzyme A synthase | Neurodegeneration with brain iron accumulation 6, pontocerebellar hypoplasia, type 12 |
| chr3:143567048 | C>T | **SLC9A9** | D007 | 3 | solute carrier family 9 member A9 | Autism Susceptibility AUTS 16 |
| chr6:31827960 | G>A | **NEU1** | D008 | 3 | neuraminidase 1 | Neuraminidase Deficiency |
| chr11:66029625 | T>C | **KLC2** | D009 | 3 | kinesin light chain 2 | Spastic Paraplegia, Optic Atrophy, and Neuropathy; SPOAN |
| chr5:178581083 | C>T | **ADAMTS2** | D010 | 3 | ADAM metalloproteinase with thrombospondin type 1 motif 2 | Ehlers-Danlos syndrome, dermatosparaxis type |
| chr6:43018723 | C>T | **CUL7** | D013 | 3 | cullin 7 | Three M syndrome 1 |
| chr19:50826985 | C>T | **KCNC3** | D013 | 3 | potassium voltage-gated channel subfamily C member 3 | Spinocerebellar ataxia 13; SCA13 |

### Overrepresentation analysis

Twenty of these 79 genes (25%) are implicated in monogenic neurodevelopmental syndromes, with both dominant and recessive modes of inheritance in the Online Mendelian Inheritance in Man (OMIM) database. We performed an overrepresentation analysis of our gene-list on a list of 2450 genes having a Central Nervous System (CNS) phenotype annotation in OMIM clinical synopsis (OMIM-CNS) among 20,203 protein coding genes in the human genome (Supplementary Table 4). The prioritized gene list from multi-sample F-SMI was significantly enriched for genes implicated in monogenic CNS syndromes (Fisher’s Exact test OR = 2.47, 95%CI = 1.41 – 4.17, p = 0.001). These gene-phenotype relationships along with the genomic coordinates and pathogenicity prediction of the identified variants in this sample are described in Table 2.

To check if the observed overrepresentation was specific to the CNS, we derived 19 additional gene-lists that encompassed genes implicated in OMIM syndromes affecting other organ systems (e.g. ‘cs_head_and_neck_head’, ‘cs_cardiovascular’ ‘cs_respiratory’ etc.). Among the 20 OMIM derived gene-lists (CNS + above 19), statistically significant overrepresentation at p < 0.0025 (0.05/20) was noted for gene lists annotated with clinical synopsis terms ‘central nervous system’ (p_cor_ = 0.027), ‘head and neck’ (p_cor_ = 0.042), and a nominally significant overrepresentation for ‘peripheral nervous system (0.057) (Supplement S2 and S3 and Supplementary Table 4) suggesting that the prioritized genes were specifically overrepresented in clinical conditions involving these systems.

To confirm whether the observed overrepresentation is truly related to the SMI under investigation, we performed an overrepresentation analysis in a set of genes (n = 918) carrying RPDs in the 60 unrelated controls. While this number was statistically significant (OR = 1.71, 95%CI = 1.43 to 2.03), the magnitude of effect was lower when compared to the effect noted for the set of 79 genes prioritized in within-pedigree analysis (OR = 2.47, 95%CI = 1.41 to 4.17, one tailed p-value = 0.089). Furthermore, the observed overrepresentation for control gene-set harboring RPDs was nonspecific as significant effects were noted for 16 out of 20 OMIM clinical synopsis genes-lists.

As the unrelated controls were from distinct nuclear pedigrees, we also compared the effect size of the overrepresentation analysis statistic between genes harboring ‘segregating’ and ‘non-segregating’ RPDs among the cases within affected pedigrees. The magnitude of effect for overrepresentation was lower in the ‘non-segregating’ RPD gene set compared to segregating gene set (supplementary section S3, supplementary figure 1).

Unlike the OMIM-CNS gene-list, we did not note an overrepresentation of the synaptic gene-list (1233 genes) from SynGO database ^31^ for the set of 79 genes with variants segregating within pedigrees (Fisher’s Exact test OR = 1.3, 95%CI = 0.58 – 3.26, p = 0.34).

### Clinical significance of prioritized genes

Variants in 20 prioritized OMIM-CNS genes were noted in 14 of the 16 multi-sample F-SMI. Clinical features that overlapped with those described in the primary OMIM syndrome were noted in members of four of these families (Table 3, Figure 1).

**Table 3:** Partial expression of symptoms from OMIM syndrome within an SMI family.

| Family Number | Gene | OMIM Clinical Syndrome | Partial overlaps in clinical features in WES sample* |
| --- | --- | --- | --- |
| D007 | COASY (OMIM 615643, 618266) | Neurodegeneration with Brain Iron Accumulation 6 (Dystonia, Parkinsonism, Cognitive Decline) | Dystonia with low dose antipsychotic, Tremors |
|  | ANTXR1 (OMIM 230740) | Growth Retardation, Alopecia, Pseudoanodontia and Optic Atrophy (IDD, Delayed motor development) | Intellectual disability |
| D012 | GEMIN5 (OMIM 619333) | Neurodevelopmental Disorder with Cerebellar Atrophy and Motor Dysfunction (Global developmental delay, IDD, speech Delay) | Learning disability<br>Marfanoid features |
| D002 | JAG1 (OMIM 118450) | Alagille Syndrome 1 (Absent deep tendon reflexes) | Delayed deep tendon reflexes |
| D006 | SCN3A (OMIM 617935, 617938) | Epilepsy Familial Focal with Variable Foci 4 (Learning disability, Speech delay) | Tardive dystonia<br>Coarse tremors<br>Academic difficulties |
|  |  | Developmental Epileptic Encephalopathy 62 (GDD, hyperkinetic movements) | Tardive dystonia<br>Coarse tremors |
|  | SOX5 (OMIM 616803) | Lamb Shaffer Syndrome (IDD, Speech delay) | Academic difficulties |
|  | DNA2 (OMIM 615807) | Seckel Syndrome (GDD, IDD) | Academic Difficulties |
| IDD - intellectual or developmental disability, GDD – Global developmental delay |  |  |  |
| * Partial symptom overlap was noted in a single family member in each of these families. |  |  |  |

### Functional significance of prioritized genes

These 79 prioritized genes in the within-pedigree analysis were significantly more conserved, in comparison with the background list of 20124 remaining protein coding genes (mean (SD) conservation score (Zoonomia ^32^) 0.63 (0.13) vs 0.59 (0.18), p =0.014). Among these genes, 20 OMIM-CNS genes were significantly more mutation intolerant as compared to the other 59 genes, as suggested by lower LOEUF score ^33^ ((mean (SD) - 0.6 (0.32) vs 0.99 (0.48), p = 0.001)) and a higher pLI score ^34^ ((mean (SD) - 0.41 (0.49) vs 0.16 (0.36), p = 0.05)). The mean expression of these 20 genes was also significantly higher compared to the other 59 genes in the brain cortex (mean (SD) -15.9 (15.3) vs 5.2 (7.5), p = 0.006); and rest of the brain (mean (SD) -16.9 (15.2) vs 6.02 (8.4), p = 0.005).

### Across pedigree association analysis using Sequence Kernel Association Test (SKAT)

We performed a gene-set-wise association analysis for higher burden of RPD variants in a synaptic gene-set (1233 genes) from SynGO database ^31^ and the OMIM-CNS gene-set (2450 genes). In the SKAT analysis accounting for kinship between cases and unrelated-controls, a statistically significant association was noted for synaptic gene-set (case burden = 153, unrelated-control burden = 41, SKAT-p = 0.016). The association for synaptic gene-set was significant in a sensitivity analysis employing 10*6 permutation tests on SKAT. The association for OMIM CNS gene-set, however, was not statistically significant (SKAT-p = 0.29).

We performed a preliminary gene-wise association analysis to examine genes with higher burden of RPD variants compared to controls (within the constraints of the study sample). We noted a significant association in gene-wise SKAT surpassing genome wide significance threshold (2.4E-6) for *CTBP2* gene and nominally significant associations surviving FDR correction for *ZNF717, SLC9B1, POTEE, CISD2 ABCD1, and DEFB108B* genes (Table 4, Supplementary table 05).

**Table 4:** Gene-set and gene-wide across pedigree association analysis.

| Gene-set wise burden |  |  |  |  |  |
| --- | --- | --- | --- | --- | --- |
| Gene | Variant count | case burden | FC burden | UC burden | SKAT p value |
| Synaptic genes | 187 | 153 | 59 | 41 | 0.014 |
| OMIM CNS genes | 569 | 148 | 60 | 59 | 0.19 |
| Gene wise burden |  |  |  |  |  |
| Gene | Variant count | case burden | FC burden | UC burden | SKAT p value |
| *CTBP2* | 3 | 136 | 39 | 2 | <0.000001 |
| *ZNF717* | 3 | 82 | 44 | 15 | 0.000121 |
| *SLC9B1* | 2 | 85 | 59 | 10 | 0.000448 |
| *POTEE* | 3 | 62 | 47 | 10 | 0.000627 |
| *ABCD1* | 4 | 51 | 31 | 7 | 0.013 |
| *CISD2* | 1 | 29 | 44 | 1 | 0.025 |
| *DEFB108B* | 1 | 47 | 37 | 8 | 0.047 |
| FC – Family control, UC – Unrelated control, SKAT – sequence kernel association test |  |  |  |  |  |

## Discussion

Employing WES in families with multiple members detected to have SMIs, we identify segregation of rare deleterious variants in several genes. A significant proportion of these genes are implicated in neuropsychiatric syndromes with Mendelian inheritance, such as *(GEMIN5, COASY, CACNA1B* etc, see Table 2). Majority (78/79) of the prioritized variants were noted to be private to fourteen multi-sample pedigrees. In four of these families, review of health records revealed evidence of clinical features that overlapped with the primary OMIM syndrome. The 79 genes prioritized by pedigree analysis tended to be more conserved, and more intolerant of variation, when compared to the remaining set of 20124 genes. In addition, in the across-pedigree association analysis, cases harbored an increased burden of rare deleterious variants in genes involved in the structural and functional integrity of synapses, as compared to unrelated controls.

The overrepresentation of genes in within-pedigree prioritization was largely specific to disorders of ‘central nervous system’ (CNS), as genes with a clinical synopsis annotation to this category showed the strongest overrepresentation. Among 19 additional clinical synopsis categories, significant overrepresentation was noted for ‘head and neck’ term. This overlap with ‘head and neck’ may be explained by tightly interlinked developmental and molecular processes that regulate cranio-facial and brain development ^35^. While there was evidence for increased burden of RPDs in genes relevant to CNS, we did not find a system-specific enrichment for genes harboring RPDs among 60 unrelated controls or the genes harboring non-segregating RPDs.

Among the 20 variants in genes implicated in CNS syndromes, three variants had been previously reported in the ClinVar database in the context of the primary Mendelian syndrome (Table 3). Of these, the variants in *ADAMTS2* and *JAG1* were noted to be of ‘uncertain significance’ and *AFG3L2* variant was recorded as ‘likely benign’ ^36^. The remaining 17 variants have not been previously reported in the ClinVar database, in the context of a primary Mendelian syndrome. Interestingly, a duplication at the same site where we find a G allele insertion (Chr21: 47860904) in the *PCNT* gene (implicated in Microcephalic osteodysplastic primordial dwarfism type II), has been identified as a pathogenic variant in ClinVar, suggesting the significance of this site in protein function.

Pleiotropic influence for *de novo* variants and genes between SCZ, BD, autism and neurodevelopmental syndromes has been reported in large trio-based studies ^7,18,23^. Our analysis also suggests genetic pleiotropy between Mendelian monogenic syndromes, and complex SMI syndromes. Ten families with variants in genes implicated in Mendelian neuropsychiatric syndrome did not have the ‘classical’ features of the primary syndrome. However, in four of these families we noted partial overlaps in clinical features, in some of the individuals with SMI (Table 3). This may suggest incomplete penetrance and expression of the primary syndrome, and possible pleiotropic expression of a psychiatric syndrome for identified gene variants.

To examine if the observed pleiotropy between monogenic and complex SMI was specific to the current sample structure, population, or variant/gene prioritization approach we curated a list of 142 genes identified in 10 recent NGS studies in SCZ, BD, OCD, AD in the literature (Supplementary Table 06). With 37 of 142 (26.1%) genes implicated in a Mendelian syndrome, we noted a statistically significant overrepresentation (Fisher’s Exact test OR = 2.29, p < 0.0001). Despite heterogeneity across these 10 studies with respect to the psychiatric syndrome, sample selection and variant prioritization methods, consistent evidence of overlap of risk between Mendelian and complex disease, at the level of aggregate list of genes harboring rare variants is noted.

Thus, while certain mutations in Mendelian disease genes result in early onset severe neurodevelopmental phenotype, other variants in the same genes may result in more subtle anatomical and functional consequences, that perhaps predispose to late-onset neuropsychiatric disorders ^37^. These phenotype level effects of a given mutation, in a given gene, may further be moderated by background genetic effects ^38,39^ and the role of the gene in neurodevelopment, and plasticity, over time ^40^. Lastly, behavioral symptoms are frequently encountered with many Mendelian neurodevelopmental syndromes, and familial SMI could represent a cumulative consequence of multiple rare variants in more than one gene ^41^.

We noted a higher burden of RPD variants in genes related to synaptic function, in the across-pedigree association analysis using SKAT. Both family-based and population-based genetic studies probing common and rare variants across a spectrum of SMI syndromes have implicated multiple synaptic genes with consequences on the structure and function of synapse ^18,42-44^. As in the case-control WES study in South African Xhosa ^4^, and using the same target list of genes, we observed rare predicted-deleterious variants in 104 genes related to synaptic structure and function. Of these, variants in genes represented by the GO component ‘integral component of presynaptic membrane’ (17/104) and ‘integral component of postsynaptic membrane’ (16/104), contributed the greatest proportion. Most (95%) of the rare-predicted deleterious variants in synaptic genes were private to a pedigree, or an individual, with only 6 genes having variants across two or more pedigrees (Supplementary table 07).

In the gene-wise association test using SKAT in the cross-pedigree analysis, the signal noted at CTBP2 gene passed the threshold for genome-wide significance (p < 2.4E-6). Nominally significant associations were noted for six additional genes (Table 4). CTBP2 gene codes for C-terminal binding protein 2, an isoform of which is a major component of synaptic ribbons, and it also plays a critical role in regulating cell migration during neocortical development ^45^. The *ZNF717* gene encodes a KRAB-zinc finger transcription factor that may be critical for primate cortical evolution, specifically differentiating human cortical transcription factor expression from that of chimpanzees ^46^. Post-zygotic mutations in *ZNF717* have also been proposed to explain the discordance for expression of schizophrenia among monozygotic twins ^47^. The *SLC9B1* gene is within the cis regulatory region of a genome wide significant association locus in a recently published GWAS of schizophrenia ^48,49^. A significant association with a *CISD2* gene locus was identified in a case-case GWAS of SCZ and autism ^50^. The *ABCD1* gene on chromosome X encodes peroxisomal membrane protein called adrenoleukodystrophy protein involved in the transport of very long chain fatty acids. More than 650 mutations in this gene have been implicated in adrenoleukodystrophy syndrome, with highly variable clinical manifestations that often include cognitive and neuropsychiatric symptoms ^51^. In summary, four of the six nominally significant genes with a higher variant burden among cases had evidence in the literature with relevance to SMI and neurodevelopment.

Some limitations are to be considered while interpreting the results of this study. While each F-SMI pedigree had multiple members affected with SMI, we could sample 3 or more affected individuals in only 16 such pedigrees. Hence the within-pedigree segregation analysis could only be performed in this subsample, reducing the power of this analysis to detect additional disease relevant variants. Augmenting sampling in remaining pedigrees may yield larger number of SMI relevant signals. We noted an overrepresentation of OMIM-CNS genes but not the synaptic genes in the list of genes prioritized in within pedigree approach. In contrast, in the across pedigree case-control association analysis approach we noted an increased burden of RPD variants in synaptic genes but not in OMIM_CNS gene list. These results lead us to speculate that in the context of genetic risk for complex neuropsychiatric syndromes in familial context, two-fold burden of effects from neurodevelopmental and synaptic function genes may contribute to final disease expression.

Alignment and variant calling were performed with hg19, an earlier version of reference sequence for the human genome. A recent study has demonstrated that 0.9% of exome targets and up to 206 genes may fall in regions that are susceptible to discrepancies in the reference assemblies between hg19 and hg38 ^52^. We verified the prioritized genes in within-family segregation and across family association analysis and observed that none of these overlapped with the discrepant genes or regions ^52^. While our sample was adequately powered to confirm across pedigree association in pre-identified gene sets, the novel gene-wise associations identified in this analysis are tentative and would need confirmation in larger, diverse samples.

## Conclusion

In this F-SMI WES study, we identify several private, rare, protein altering variants that segregate among the cases within a pedigree. These variants are overrepresented among genes implicated in monogenic forms of Mendelian neuropsychiatric syndromes, suggesting pleotropic influences in neurodevelopment and functioning. We also note a greater frequency of variants in genes involved in structural and functional integrity of synapses in cases compared to controls. The study demonstrates the usefulness of NGS approaches in F-SMI to identify disease relevant variants. Future studies, involving a larger number of families, with multiple affected members, and across diverse populations, may help us explore the contribution of rare coding variants to F-SMI. Validation of the functional impact of identified variants using cell models, combined with the application of *in silico* approaches to model protein structure alterations, and interactions, may help understand the convergent and divergent developmental mechanisms that underlie rare variants and risk of complex SMI.

## Methods

### Ethics statement

The study protocol was approved by the Institutional Ethics Committee of National Institute of Mental Health and Neurosciences, Bengaluru, India, and the study procedures conformed to the provisions of the Declaration of Helsinki. All participants provided written informed consent.

### Sample

The families with multiple members with SMI (F-SMI) were identified as part of ADBS. This program is aimed at characterization of clinical and neurobiological phenotypes for neuropsychiatric syndromes and identification of genetic and molecular correlates of disease using cell models ^2,30,53^. The diagnosis of SMI was established by independent clinical evaluation by two psychiatrists based on ICD – 10 criteria ^54^. Diagnosis and current and lifetime comorbidity were further evaluated and confirmed with Mini International Neuropsychiatric Inventory 5.0.0 ^55^. The clinical status of the members belonging to the first two generations of all recruited participants was confirmed using Family Interview for Genetic Studies and pedigree charting ^56^.

Cases were individuals with a diagnosis of SMI and family controls were unaffected individuals from the same families without a lifetime diagnosis of SMI or related syndromes. In addition, we identified unaffected unrelated individuals without family history of SMI, as ‘unrelated-controls’ from the population. Families with WES data from ≥ 3 cases were identified as ‘multi-sample F-SMI’ and selected for analysis of within-family segregation of variants as described below.

### Sequencing, alignment, variant calling and quality assessment

The protocols for sequencing, alignment and variant calling including the quality controls (QC) have been previously published ^22^ (supplement section S4). In brief, Illumina Nextera exome enrichment kits targeting 62.08 Mb of human genome were used for library preparation and the Illumina Hiseq platform was used for 100 base paired-end sequencing. Raw-read QC was performed with FastQC.0.10.1 and low-quality reads (<Q20) were excluded. Alignment to human genome build hg19/GRCH37 was performed using BWA (v-0.5.9). Realignment was performed with 1000G Phase1 INDELs using GATK (v-3.6) for removal of PCR duplicates and alignment artifacts. Single Nucleotide Polymorphisms (SNPs) and short insertion deletions (INDEL) variants were called with standard parameters (min coverage = 8, MAF ≥ 0.25 and P ≤ 0.001) using Varscan2 to generate sample-wise VCFs.

### Annotation

VCFs were annotated with ANNOVAR ^57^ tool for gene, region and filter based annotation options. Variant frequencies were obtained from the gnomAD - South Asian subset (N=15,308). For *in silico* prediction of deleteriousness of coding variants, five functional annotations (*SIFT* ^58^, *LRT* ^59^, *MutationTaster* ^60^, *MutationAssessor* ^61^ and *MetaSVM* ^62^) were used.

### Variant prioritization

Variants were prioritized based on rarity (gnomAD SAS minor allele frequency ≤ 0.001) and a ‘predicted-deleterious’ functional consequence. Predicted-deleteriousness of exonic Single nucleotide variants (SNVs) were defined as protein truncating variants (stop-gain, stop-loss or start loss) or missense variants predicted to have deleterious consequence in ≥ 4 of the 5 functional prediction annotation tools. Protein truncating or out-of-frame small insertion-deletions (indels) were similarly prioritized. All subsequent analysis involved the prioritized rare predicted-deleterious (RPD) variants. Minor allele frequencies of the prioritized variants were examined in remaining populations in gnomAD database to exclude rare variants specific to South Asian sample.

### Analysis approach

To identify the RPDs that are putatively relevant to SMI syndromes we adopted two independent analytical approaches; a within-family prioritization of variants segregating with SMI and a cross-pedigree case-control association analysis.

#### A. Within-family variant selection

In the subsample of ‘multi-sample F-SMI’ with ≥3 cases, we examined RPD variants that were shared by ≥3 cases and absent in the unrelated-controls. These pedigrees are described in supplement section S5. The resulting lists of variants and the genes harboring these variants were queried with RPDs noted in the cases in previously excluded families (<3 samples from cases/family) to examine potential variant and gene level overlaps in SMI associated genes.

#### B. Case control association analysis

We performed gene-set and gene-level case-control association analysis between the entire sample of cases and unrelated-controls, after accounting for the kinship structure within the sample using Sequence Kernel Association Test (SKAT) ^63^ as implemented in the R package SKAT ^64^. Initially, the SKAT_null_emmax() function was used to approximately adjust for the overall genomic correlation among the individuals by fitting a null model for the binary case/control status incorporating the kinship matrix. The residuals thus obtained were permuted to obtain resampled residuals using SKAT_Null_Model() function. Finally, association of RPD variants with case/control status was examined using SKAT() function. The analysis was run using up to 10e+6 permutations per variant to adjust the accuracy of the p values. Gene-set wide analysis p values were adjusted for Bonferroni correction for the two gene-sets tested. Gene-based p values were corrected for multiple testing using a False Discovery Rate cut off of 5% using the BH [Benjamini Hochberg, 1995] procedure. Additionally, a raw p value threshold of 2.4E-6 was considered genome wide significant for gene-level analysis.

### Clinical and functional significance of the prioritized genes

The set of genes from within-family prioritization was examined for enrichment in genes implicated in monogenic neurodevelopmental syndromes in Online Mendelian Inheritance in Man (OMIM) database (OMIM-CNS genes) (Supplement S2) using the two tailed Fisher’s Exact test. The specificity of this enrichment to the CNS was verified by examining enrichment for 19 non-CNS gene lists among additional OMIM clinical synopsis categories (Supplement S2) and by repeating these analysis in genes harboring RPD variants in unrelated controls. In families where RPD variants in OMIM genes for particular monogenic CNS syndromes were noted, we examined the medical records for any overlapping OMIM-like clinical features. We further examined if the prioritized genes were overrepresented for synaptic genes derived from SynGO database ^31^.

We examined the mutational constraint, tissue expression and evolutionary conservation using publicly available datasets. Mutational constraint was assessed by gnomAD LOEUF ^33^ and ExAC pLI scores ^34^. Brain and cortical expression were examined using the GTEx v8 with mean expression values per gene. Conservation scores derived from 240 species alignment were adopted from the Zoonomia consortium (Zoonomia fraction of CDS phyloP ≥ 2.270 (fdr 0.05)) ^32^. Comparisons between prioritized and the background list were performed using the Welch’s t-test.

We performed a gene-set-wide association analysis between cases and unrelated-controls, across pedigrees, using SKAT modified for gene-set analysis. In this approach, we tested two *a priori* defined gene sets: OMIM-CNS genes – to test the findings of the within-pedigree analyses and for synaptic genes from SynGO database ^31^; to compare with the results of earlier WES studies that implicated synaptic genes in SMIs.

## Data Availability

All data analysis in the present work are contained in the manuscript or uploaded as supplementary files. Raw data  are available upon reasonable request to the authors

## ADBS consortium

Naren P. Rao^1^, Janardhanan C. Narayanaswamy^1^, Palanimuthu T. Sivakumar^1^, Arun Kandasamy^1^, Muralidharan Kesavan^1^, Urvakhsh Meherwan Mehta^1^, Ganesan Venkatasubramanian^1^, John P. John^1^, Odity Mukherjee^2^, Ramakrishnan Kannan^1^, Bhupesh Mehta^1^, Thennarasu Kandavel^1^, B. Binukumar^1^, Jitender Saini^1^, Deepak Jayarajan^1^, A. Shyamsundar^1^, Sydney Moirangthem^1^, K. G. Vijay Kumar^1^, Bharath Holla^1^, Jayant Mahadevan^1^, Jagadisha Thirthalli^1^, Prabha S. Chandra^1^, Bangalore N. Gangadhar^1^, Pratima Murthy^1^, Mitradas M. Panicker^3^, Upinder S. Bhalla^3^, Sumantra Chattarji^3^, Vivek Benegal^1^, Mathew Varghese^1^, Janardhan Y. C. Reddy^1^, Padinjat Raghu^3^ and Mahendra Rao^2^, Biju Viswanath^1^, Meera Purushottam^1^, Sanjeev Jain^1^

1. National Institute of Mental Health and Neurosciences, Bangalore, India
2. Institute for Stem Cell Biology and Regenerative Medicine (InStem), Bangalore, India
3. National Center for Biological Sciences (NCBS), Bangalore, India

## Acknowledgements

The authors are grateful to all the patients, their family members and healthy volunteers who participated in the study. Financial support for the study was provided by Department of Biotechnology funded grants - BT/01/CEIB/11/VI/11/2012, entitled, “Targeted generation and interrogation of cellular models and networks in neuro-psychiatric disorders using candidate genes” and BT/PR17316/MED/31/326/2015 entitled, “Accelerator program for discovery in brain disorders using stem cells” (ADBS), Pratiksha Trust and The Institute of Stem Cells and Regenerative Medicine (InStem), Bengaluru, India.

The authors would like to thank the sequencing core facility at the Institute of Genomics and Integrative Biology (IGIB), Delhi (Dr. Faruq Mohammed) and the National Centre for Biological Sciences (NCBS), Bengaluru (Dr. Awadhesh Pandit) for sample processing and WES data generation.

The authors would like to thank all investigators of ADBS consortia for providing valuable inputs to the manuscript and having final approval of the manuscript. Suhas Ganesh is affiliated with Schizophrenia Neuropharmacology Research Group at Yale university and is supported by a NARSAD young investigator grant from the Brain and Behavior Research Foundation. Biju Viswanath is funded by the Intermediate (Clinical and Public Health) Fellowship (IA/CPHI/20/1/505266) of the DBT/Wellcome Trust India Alliance.

## Additional information - Disclosure statement

The authors declare that there are no conflicts of interest with the work presented in the manuscript.

## Author contributions

SJ, MP, BV and ADBS consortium conceived the study. SG, AV, KM curated the WES data and performed analysis. DI, KN, RKN curated clinical data and assisted with the WES analysis. SB provided inputs on statistical analysis and interpretation. PFS provided inputs on bioinformatic analysis and interpretation. All authors contributed to writing, revising, and finalizing the manuscript draft.

## Supplementary material

S1 Overview of recent sequencing studies in Serious Mental Illnesses (SMI)

S2 Derivation of OMIM lists from clinical synopsis terms

S3 OMIM overrepresentation – specificity to CNS

S4 Sequencing and variant quality assessment with IGV and read depth

S5 Pedigrees included in the within pedigree prioritization

## Supplementary tables

Supplementary table 01: Within pedigree prioritization in multi-sample (≥3) families with SMI – list of 79 prioritized variants and genes

Supplementary table 02: Gene level overlaps for prioritized genes in less sample dense (≤2) pedigrees

Supplementary table 03: Variant level overlaps for prioritized genes in less sample dense (≤2) pedigrees

Supplementary table 04: Specificity of OMIM overrepresentation to Central Nervous System impacting genes

Supplementary table 05: Association results for gene-wise sequence kernel association test

Supplementary table 06: OMIM syndrome and complex SMI gene level overlap in 10 NGS studies in literature

Supplementary table 07: Gene ontology description of variant burden in synaptic genes

## References

1 Huang, M.-H. et al. Familial coaggregation of major psychiatric disorders among first-degree relatives of patients with obsessive-compulsive disorder: a nationwide study. Psychological Medicine 51, 680–687, doi:10.1017/S0033291719003696 (2020).

2 Sreeraj, V. S. et al. Psychiatric symptoms and syndromes transcending diagnostic boundaries in Indian multiplex families: The cohort of ADBS study. Psychiatry Res 296, 113647, doi:10.1016/j.psychres.2020.113647 (2021).

3 Glahn, D. C. et al. Rediscovering the value of families for psychiatric genetics research. Mol Psychiatry 24, 523–535, doi:10.1038/s41380-018-0073-x (2019).

4 Gulsuner, S. et al. Genetics of schizophrenia in the South African Xhosa. Science 367, 569–573, doi:10.1126/science.aay8833 (2020).

5 Singh, T. et al. Rare coding variants in ten genes confer substantial risk for schizophrenia. Nature 604, 509–516, doi:10.1038/s41586-022-04556-w (2022).

6 Kato, T. Whole genome/exome sequencing in mood and psychotic disorders. Psychiatry Clin Neurosci 69, 65–76, doi:10.1111/pcn.12247 (2015).

7 Howrigan, D. P. et al. Exome sequencing in schizophrenia-affected parent-offspring trios reveals risk conferred by protein-coding de novo mutations. Nat Neurosci 23, 185–193, doi:10.1038/s41593-019-0564-3 (2020).

8 Legge, S. E. et al. Genetic architecture of schizophrenia: a review of major advancements. Psychol Med, 1–10, doi:10.1017/S0033291720005334 (2021).

9 Li, M. et al. Novel genetic susceptibility loci identified by family based whole exome sequencing in Han Chinese schizophrenia patients. Transl Psychiatry 10, 5, doi:10.1038/s41398-020-0708-y (2020).

10 Forstner, A. J. et al. Whole-exome sequencing of 81 individuals from 27 multiply affected bipolar disorder families. Transl Psychiatry 10, 57, doi:10.1038/s41398-020-0732-y (2020).

11 Goes, F. S. et al. Exome Sequencing of Familial Bipolar Disorder. JAMA Psychiatry 73, 590–597, doi:10.1001/jamapsychiatry.2016.0251 (2016).

12 Sul, J. H. et al. Contribution of common and rare variants to bipolar disorder susceptibility in extended pedigrees from population isolates. Transl Psychiatry 10, 74, doi:10.1038/s41398-020-0758-1 (2020).

13 Toma, C. et al. An examination of multiple classes of rare variants in extended families with bipolar disorder. Transl Psychiatry 8, 65, doi:10.1038/s41398-018-0113-y (2018).

14 Halvorsen, M. et al. Exome sequencing in obsessive-compulsive disorder reveals a burden of rare damaging coding variants. Nat Neurosci 24, 1071–1076, doi:10.1038/s41593-021-00876-8 (2021).

15 Jia, X. et al. Investigating rare pathogenic/likely pathogenic exonic variation in bipolar disorder. Mol Psychiatry, doi:10.1038/s41380-020-01006-9 (2021).

16 Palmer, D. S. et al. Exome sequencing in bipolar disorder reveals shared risk gene *AKAP11* with schizophrenia. medRxiv, 2021.2003.2009.21252930, doi:10.1101/2021.03.09.21252930 (2021).

17 Ganna, A. et al. Quantifying the Impact of Rare and Ultra-rare Coding Variation across the Phenotypic Spectrum. Am J Hum Genet 102, 1204–1211, doi:10.1016/j.ajhg.2018.05.002 (2018).

18 Nishioka, M. et al. Systematic analysis of exonic germline and postzygotic de novo mutations in bipolar disorder. Nat Commun 12, 3750, doi:10.1038/s41467-021-23453-w (2021).

19 Fromer, M. et al. De novo mutations in schizophrenia implicate synaptic networks. Nature 506, 179–184, doi:10.1038/nature12929 (2014).

20 Genovese, G. et al. Increased burden of ultra-rare protein-altering variants among 4,877 individuals with schizophrenia. Nat Neurosci 19, 1433–1441, doi:10.1038/nn.4402 (2016).

21 Kenny, E. M. et al. Excess of rare novel loss-of-function variants in synaptic genes in schizophrenia and autism spectrum disorders. Mol Psychiatry 19, 872–879, doi:10.1038/mp.2013.127 (2014).

22 Ganesh, S. et al. Exome sequencing in families with severe mental illness identifies novel and rare variants in genes implicated in Mendelian neuropsychiatric syndromes. Psychiatry Clin Neurosci 73, 11–19, doi:10.1111/pcn.12788 (2019).

23 Rees, E. et al. Schizophrenia, autism spectrum disorders and developmental disorders share specific disruptive coding mutations. Nat Commun 12, 5353, doi:10.1038/s41467-021-25532-4 (2021).

24 Brainstorm-Consortium et al. Analysis of shared heritability in common disorders of the brain. Science 360, doi:10.1126/science.aap8757 (2018).

25 Cristino, A. S. et al. Neurodevelopmental and neuropsychiatric disorders represent an interconnected molecular system. Mol Psychiatry 19, 294–301, doi:10.1038/mp.2013.16 (2014).

26 Grotzinger, A. D. Shared genetic architecture across psychiatric disorders. Psychol Med, 1–7, doi:10.1017/S0033291721000829 (2021).

27 Gandal, M. J. et al. Shared molecular neuropathology across major psychiatric disorders parallels polygenic overlap. Science 359, 693–697, doi:10.1126/science.aad6469 (2018).

28 Radonjic, N. V. et al. Structural brain imaging studies offer clues about the effects of the shared genetic etiology among neuropsychiatric disorders. Mol Psychiatry 26, 2101–2110, doi:10.1038/s41380-020-01002-z (2021).

29 Grotzinger, A. D. et al. Genetic architecture of 11 major psychiatric disorders at biobehavioral, functional genomic and molecular genetic levels of analysis. Nat Genet 54, 548–559, doi:10.1038/s41588-022-01057-4 (2022).

30 Viswanath, B. et al. Discovery biology of neuropsychiatric syndromes (DBNS): a center for integrating clinical medicine and basic science. BMC Psychiatry 18, 106, doi:10.1186/s12888-018-1674-2 (2018).

31 Koopmans, F. et al. SynGO: An Evidence-Based, Expert-Curated Knowledge Base for the Synapse. Neuron 103, 217–234 e214, doi:10.1016/j.neuron.2019.05.002 (2019).

32 Genereux, D. P. et al. A comparative genomics multitool for scientific discovery and conservation. Nature 587, 240–245, doi:10.1038/s41586-020-2876-6 (2020).

33 Karczewski, K. J. et al. The mutational constraint spectrum quantified from variation in 141,456 humans. Nature 581, 434–443, doi:10.1038/s41586-020-2308-7 (2020).

34 Fuller, Z. L., Berg, J. J., Mostafavi, H., Sella, G. & Przeworski, M. Measuring intolerance to mutation in human genetics. Nat Genet 51, 772–776, doi:10.1038/s41588-019-0383-1 (2019).

35 Naqvi, S. et al. Shared heritability of human face and brain shape. Nat Genet 53, 830–839, doi:10.1038/s41588-021-00827-w (2021).

36 Richards, S. et al. Standards and guidelines for the interpretation of sequence variants: a joint consensus recommendation of the American College of Medical Genetics and Genomics and the Association for Molecular Pathology. Genet Med 17, 405–424, doi:10.1038/gim.2015.30 (2015).

37 Zhu, X., Need, A. C., Petrovski, S. & Goldstein, D. B. One gene, many neuropsychiatric disorders: lessons from Mendelian diseases. Nat Neurosci 17, 773–781, doi:10.1038/nn.3713 (2014).

38 Chen, R. et al. Analysis of 589,306 genomes identifies individuals resilient to severe Mendelian childhood diseases. Nat Biotechnol 34, 531–538, doi:10.1038/nbt.3514 (2016).

39 Riordan, J. D. & Nadeau, J. H. From Peas to Disease: Modifier Genes, Network Resilience, and the Genetics of Health. Am J Hum Genet 101, 177–191, doi:10.1016/j.ajhg.2017.06.004 (2017).

40 Ball, G. et al. Cortical morphology at birth reflects spatiotemporal patterns of gene expression in the fetal human brain. PLoS Biol 18, e3000976, doi:10.1371/journal.pbio.3000976 (2020).

41 Wang, Y. C. et al. Identification of ultra-rare missense mutations associated with familial schizophrenia by whole-exome sequencing. Schizophr Res 235, 60–62, doi:10.1016/j.schres.2021.07.027 (2021).

42 Forrest, M. P., Parnell, E. & Penzes, P. Dendritic structural plasticity and neuropsychiatric disease. Nat Rev Neurosci 19, 215–234, doi:10.1038/nrn.2018.16 (2018).

43 Hall, J., Trent, S., Thomas, K. L., O’Donovan, M. C. & Owen, M. J. Genetic risk for schizophrenia: convergence on synaptic pathways involved in plasticity. Biol Psychiatry 77, 52–58, doi:10.1016/j.biopsych.2014.07.011 (2015).

44 Taoufik, E., Kouroupi, G., Zygogianni, O. & Matsas, R. Synaptic dysfunction in neurodegenerative and neurodevelopmental diseases: an overview of induced pluripotent stem-cell-based disease models. Open Biol 8, doi:10.1098/rsob.180138 (2018).

45 Wang, H. et al. ZEB1 Represses Neural Differentiation and Cooperates with CTBP2 to Dynamically Regulate Cell Migration during Neocortex Development. Cell Rep 27, 2335–2353 e2336, doi:10.1016/j.celrep.2019.04.081 (2019).

46 Nowick, K., Gernat, T., Almaas, E. & Stubbs, L. Differences in human and chimpanzee gene expression patterns define an evolving network of transcription factors in brain. Proc Natl Acad Sci U S A 106, 22358–22363, doi:10.1073/pnas.0911376106 (2009).

47 Castellani, C. A. et al. Post-zygotic genomic changes in glutamate and dopamine pathway genes may explain discordance of monozygotic twins for schizophrenia. Clin Transl Med 6, 43, doi:10.1186/s40169-017-0174-1 (2017).

48 Lam, M. et al. Comparative genetic architectures of schizophrenia in East Asian and European populations. Nat Genet 51, 1670–1678, doi:10.1038/s41588-019-0512-x (2019).

49 Li, Z. et al. Genome-wide association analysis identifies 30 new susceptibility loci for schizophrenia. Nat Genet 49, 1576–1583, doi:10.1038/ng.3973 (2017).

50 Peyrot, W. J. & Price, A. L. Identifying loci with different allele frequencies among cases of eight psychiatric disorders using CC-GWAS. Nat Genet 53, 445–454, doi:10.1038/s41588-021-00787-1 (2021).

51 Engelen, M. et al. X-linked adrenoleukodystrophy (X-ALD): clinical presentation and guidelines for diagnosis, follow-up and management. Orphanet J Rare Dis 7, 51, doi:10.1186/1750-1172-7-51 (2012).

52 Li, H. et al. Exome variant discrepancies due to reference-genome differences. Am J Hum Genet 108, 1239–1250, doi:10.1016/j.ajhg.2021.05.011 (2021).

53 Sreeraj, V. S. et al. Cross-diagnostic evaluation of minor physical anomalies in psychiatric disorders. J Psychiatr Res 142, 54–62, doi:10.1016/j.jpsychires.2021.07.028 (2021).

54 Who, W. H. O. The ICD-10 classification of mental and behavioural disorders: clinical descriptions and diagnostic guidelines. (World Health Organization, 1992).

55 Sheehan, D. V. et al. The Mini-International Neuropsychiatric Interview (M.I.N.I.): the development and validation of a structured diagnostic psychiatric interview for DSM-IV and ICD-10. J Clin Psychiatry 59 Suppl 20, 22–33;quiz 34-57 (1998).

56 Maxwell, M. E. (1992).

57 Wang, K., Li, M. & Hakonarson, H. ANNOVAR: functional annotation of genetic variants from high-throughput sequencing data. Nucleic Acids Res 38, e164, doi:10.1093/nar/gkq603 (2010).

58 Kumar, P., Henikoff, S. & Ng, P. C. Predicting the effects of coding non-synonymous variants on protein function using the SIFT algorithm. Nat Protoc 4, 1073–1081, doi:10.1038/nprot.2009.86 (2009).

59 Chun, S. & Fay, J. C. Identification of deleterious mutations within three human genomes. Genome Res 19, 1553–1561, doi:10.1101/gr.092619.109 (2009).

60 Schwarz, J. M., Cooper, D. N., Schuelke, M. & Seelow, D. MutationTaster2: mutation prediction for the deep-sequencing age. Nature Methods 11, 361–362, doi:10.1038/nmeth.2890 (2014).

61 Reva, B., Antipin, Y. & Sander, C. Predicting the functional impact of protein mutations: application to cancer genomics. Nucleic Acids Research 39, e118–e118, doi:10.1093/nar/gkr407 (2011).

62 Dong, C. et al. Comparison and integration of deleteriousness prediction methods for nonsynonymous SNVs in whole exome sequencing studies. Hum Mol Genet 24, 2125–2137, doi:10.1093/hmg/ddu733 (2015).

63 Lee, S. et al. Optimal unified approach for rare-variant association testing with application to small-sample case-control whole-exome sequencing studies. Am J Hum Genet 91, 224–237, doi:10.1016/j.ajhg.2012.06.007 (2012).

64 SNP-Set (Sequence) Kernel Association Test v. 2.0.1 (2020).

