## Supplement 1 for "Whole Exome Sequencing in dense families suggests genetic pleiotropy amongst Mendelian and complex neuropsychiatric syndromes"

**S1** **Overview of recent sequencing studies in Serious Mental Illnesses (SMI)**

**Family studies:** Using WES to analyze 65 families with SCZ, Li et al, identified several *de novo* mutations (DNMs) and risk alleles in genes previously implicated in SCZ as well as novel genes that contributed to risk (1). Studies suggest that rare loss-of-function variants occur more often in those with SCZ compared to control samples (2). An analysis of parent-offspring trios with SCZ suggested that DNMs were detected much more often in genes expressed in the brain, implicated in neurodevelopmental disorders, or under evolutionary constraint (3). A similar observation was also made with respect to inherited variants that contribute to risk of autism, perhaps attributable to variants that were likely to disrupt gene function, in certain pathways, suggesting a multi-hit model (4). Overlaps between neurodevelopmental disorders, autism and risk of schizophrenia are thus observed quite often, and it is suggested that they may have an overlapping pathophysiology.

A WES analysis of 81 individuals from 27 families (with several members diagnosed as BD) again suggested that several rare variants that could contribute to risk of disease. While some of these could be construed as being highly penetrant (shared across families, present in most affected within a family), others too were thought to be plausible risk factors. In addition, overlaps with rare variants implicated in SZ and autism were also identified (5). A family-based WES study in BD with 15 families noted that rare predicted pathogenic variants shared by 3 or more cases were overrepresented in genes coding for post-synaptic density proteins (6). Another family-based study noted small to moderate effect increase in the burden of rare deleterious variants in extended pedigrees with multiple affected members. This analysis was restricted to genes that were previously associated with BD. (7).

A trio-based WES study in OCD noted an increased burden of protein altering variations, which were statistically significant in the *SLITRK5* and *CHD8* genes (8). The *SLITRK* gene family in involved in synaptic regulation, while the *CHD8* has been implicated as a risk gene in BD in a case control WES study (9).

Interestingly, while most common and rare variant case-control studies in AD have implicated immune and amyloid processing genes (10), a recent family based WGS implicated rare variants in neurodevelopmental and synaptic function genes (11). In a family-based WES study with multiple affected members with SMI, we have previously demonstrated segregation of rare deleterious variants in genes implicated in Mendelian neuropsychiatric syndromes, that are private to each pedigree (12).

**Case-control studies:** A WES study comparing around ~900 SCZ cases and matched controls of South African Xhosa noted an increased burden of ultra-rare damaging variants in genes related to synaptic function, specifically those involved in glutamate, gamma aminobutyric acid, and dopamine signaling (13). The largest study to date involving 24248 SCZ cases and 97322 controls identified increased burden of large effect ultra-rare variants in 10 exome wide significant genes again implicating molecular functions involving formation, structure and function of the synapse (14). A WES study in BD with ~14000 cases and controls also noted a significant burden of ultra-rare protein truncating variants along with significant enrichment of these signals with the gene-set implicated in SCZ WES (9). However, another study that analyzed pathogenic or likely pathogenic variants across the exome in BD did not find a replicable signal for increased burden of such variants in cases compared to controls (15).

**S2 Derivation of gene-lists from clinical synopsis terms**

The Online Mendelian Inheritance in Man (OMIM) (16) is an exhaustive online compendium of known gene-phenotype relationship relationships. We used ‘clinical synopsis search fields’ in the ‘Gene map advanced search’ option in OMIM to derive lists of genes implicated in syndromes annotated with a clinical synopsis term referring to a specific organ system. For example, to search for genes implicated in syndrome affecting central nervous system we used the clinical synopsis term ‘cs_neurologic_central_nervous_system_exists’. A similar approach was used to derive gene list tables for 19 additional systems e.g. ‘cs_genitourinary_exists’ for genes involved in syndromes where genitourinary system is affected; ‘cs_head_and_neck_head_exists’ for genes involved in syndromes where head and neck development is affected. From these gene map tables, gene lists were derived by removing duplicate entries within the ‘Approved Symbol Column’. All OMIM searches for gene lists were performed on July 20, 2021. A list of 20 systems for which gene lists were generated are provided in supplementary table 4.

**S3 OMIM overrepresentation – specificity to CNS**

Over representation of the prioritized 79 genes in within pedigree segregation analysis for the 20 OMIM clinical synopsis gene lists noted above was examined using fisher’s exact test. Bonferroni correction was applied for multiple hypothesis testing. The strongest statistically significant association was noted for gene-list for ‘central nervous system’. Additionally. significant overrepresentation was also noted for gene list implicated in ‘head and neck’ syndromes (Supplementary table 4a).

To examine if the observed overrepresentation was relevant to SMIs and not due to a background propensity for higher burden of RPDs in genes relevant to CNS function derived by selection forces, we repeated the above analysis in a set of genes (n = 918) harboring RPDs among 60 unrelated controls. an overrepresentation for OMIM CNS syndromes for rare putatively deleterious variants is seen both in cases (within family segregation) and in unrelated population controls. However, for the 79 genes with RPDs segregating in ≥3 cases with SMI, the enrichment is selective for CNS and head and neck syndromes. Among the controls genes annotated with several clinical synopsis terms including laboratory abnormalities, skin hair nail, abdomen, skeletal, cardiovascular, genitourinary systems show larger magnitude of overrepresentation compared to central nervous system. This finding suggests that the genes carrying RPDs among controls show non-specific enrichment to several genes implicated in mendelian syndromes including CNS but without a specific predilection for CNS. The smaller p-values noted in the control set (4b) compared to the case set (4a) can be attributed to larger number of RPD variant harboring genes (918) among the controls.

Furthermore, we compared the magnitudes of overrepresentation for OMIM-CNS genes in 79 genes with variants segregating in SMI families (OR = 2.47, 95%CI – 1.41 to 4.17) versus genes carrying RPDs in unrelated controls (OR = 1.71, 95%CI – 1.43 to 2.03). A one-tailed test to examine if logarithm of case OR is greater than control OR using the formulae (z = δlogOR/SE(δ) resulted in z = 1.343 and a p value = 0.089, suggesting a trend for larger magnitude for overrepresentation for OMIM-CNS genes in the gene list derived from case families. Although we observe this trend, the absence of statistically significant difference may be attributed to large standard errors resulting from a small gene list for within family segregation. As most of the RPDs are private, future studies in larger number of pedigrees may provide a more robust signal.

To examine if the selection of ‘segregating’ variants among cases influence the CNS relevance of the derived gene list, we compared this list with a list of genes with ‘non-segregating’ RPDs among cases within families. The logORs and SE of logORs from these analyses are plotted in supplementary figure 1 under the section S3 and pasted below.

We note that the largest OR for overrepresentation is noted for gene list (n = 79) derived from RPDs segregating in cases. And this estimate is larger compared to gene-lists carrying RPDs in all other gene sets harboring – a. ‘non-segregating’ RPDs within pedigrees, b. RPDs in family controls, c – RPDs in unrelated controls.

Supplementary figure 1. Comparison of magnitudes of enrichment across gene-sets from familial cases, family controls and unrelated population controls

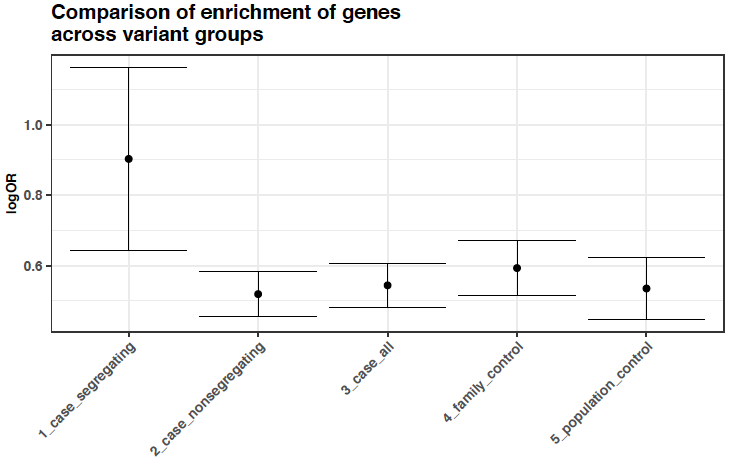

Supplementary figure 1 presents a comparison of the magnitudes of overrepresentation for OMIM-CNS genes in 1. The set of 79 genes with segregating RPD variants in SMI cases within families; 2. Genes carrying non-segregating RPD variants in SMI cases within families; 3. All genes harboring RPD variants among cases; 4. All genes harboring RPD variants among family controls; 5 All genes harboring RPD variants among unrelated population controls. Logarithm of Odds Ratios from Fisher’s test and Standard Errors of Log ORs are plotted along the y axis.

**S4 Sequencing and variant quality assessment with IGV and read depth**

Illumina Nextera exome enrichment kits targeting 62.08 Mb of human genome were used for library preparation and the Illumina Hiseq platform was used for 100 base paired-end sequencing. Raw-read QC was performed with FastQC.0.10.1 and low-quality reads (<Q20) were excluded. Alignment to human genome build hg19/GRCH37 was performed using BWA (v-0.5.9). Realignment was performed with 1000G Phase1 INDELs using GATK (v-3.6) for removal of PCR duplicates and alignment artifacts. Mean percentage of mapped reads and mapping quality scores across samples were 98.8% and 55.03 respectively. Median coverage at 20X was 62.31. For each sample, coverage and depth were further examined at each chromosome and for a sample set of 50 genes of interest. Blood group, HLA and sex matching were performed as part of QC pipeline. Single Nucleotide Polymorphisms (SNPs) and short insertion deletions (INDEL) variants were called with standard parameters (min coverage = 8, MAF ≥ 0.25 and P ≤ 0.001) using Varscan2 to generate sample-wise VCFs. For a subset of the prioritized variants, realigned BAM files were visualized in Integrative Genomics Viewer (IGV) to confirm the variant calls.

Supplementary figure 2: Viewing Read alignments using Integrative Genomics Viewer(IGV) to verify SNV positions in few samples.

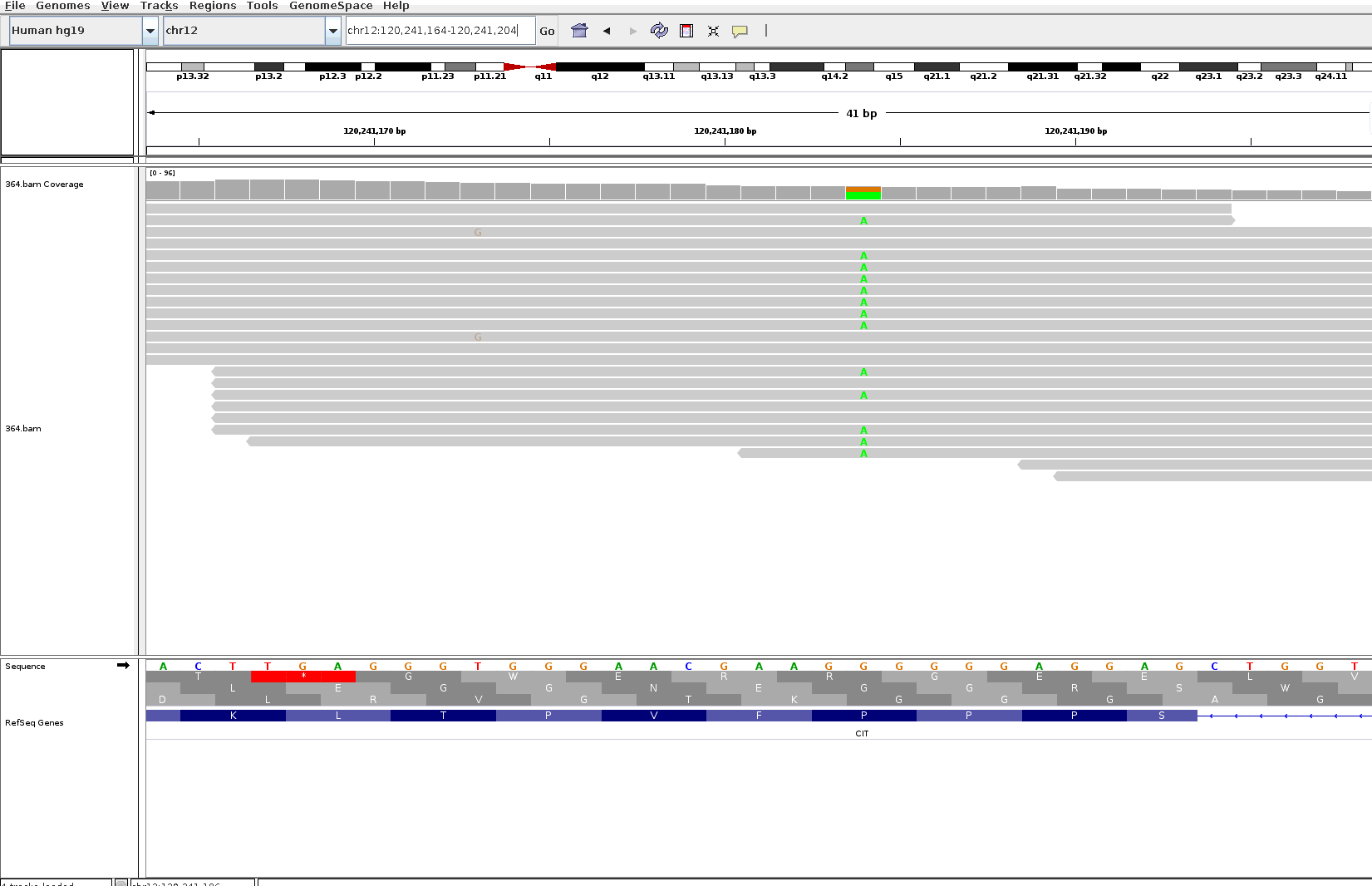

Figure 2A: The alternate allele A is present in gene CIT as a nonsynonymous SNV on chromosome 12 at position 120241184 in a sample belonging to the Family D004

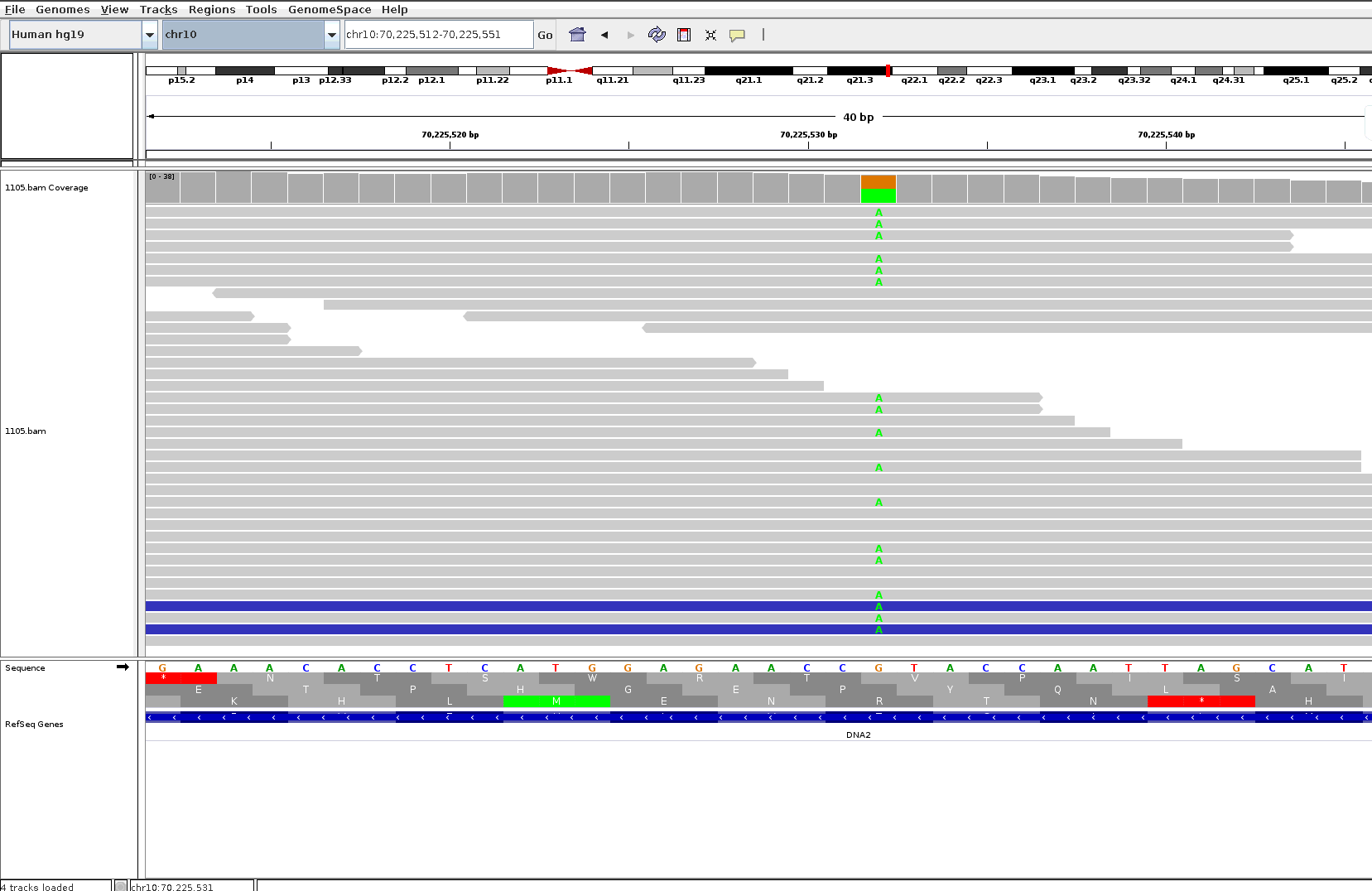

Figure 2B: The alternate allele A is present in gene DNA2 as a nonsynonymous SNV on chromosome 10 at position 70225532 in a sample belonging to the Family D006

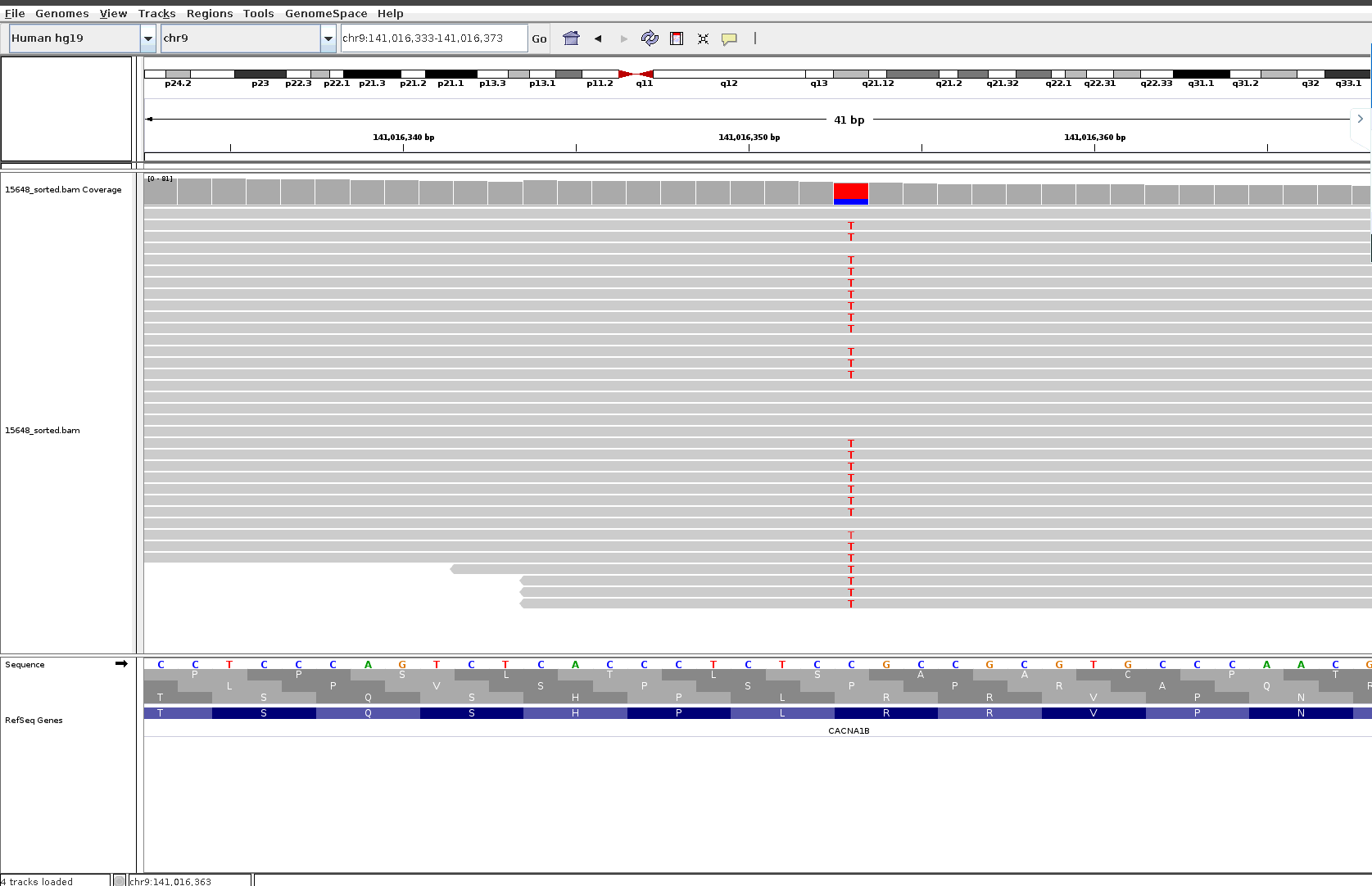

Figure 2C: The alternate allele T is present in gene CACNA1B as a nonsynonymous SNV on chromosome 9 at position 141016353 in a sample belonging to the Family D002

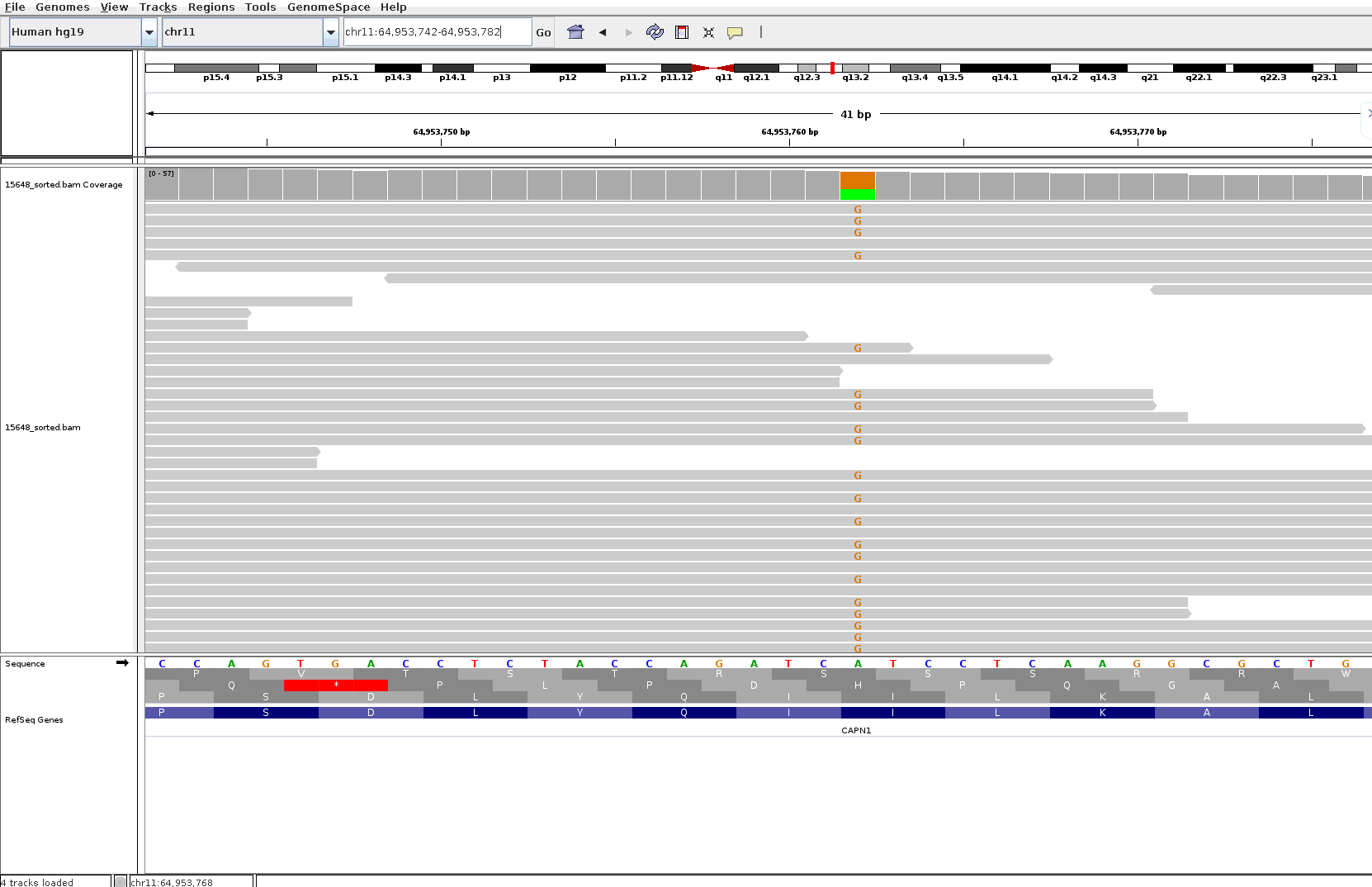

Figure 2D: The alternate allele G is present in gene CAPN1 as a nonsynonymous SNV on chromosome 11 at position 64953762 in a sample belonging to the Family D002

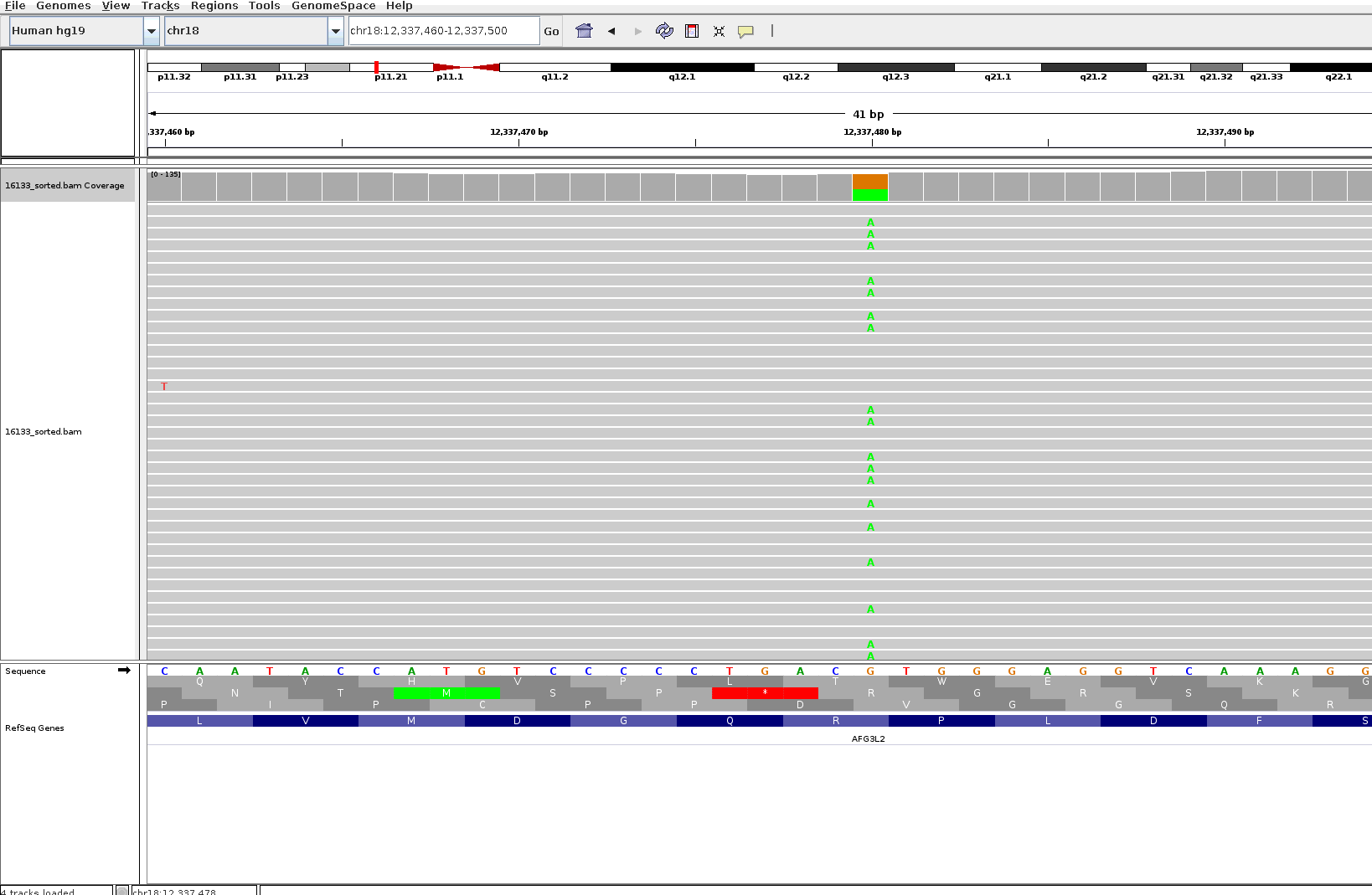

Figure 2E: The alternate allele A is present in gene AFG3L2 as a nonsynonymous SNV on chromosome 18 at position 12337480 in a sample belonging to the D002

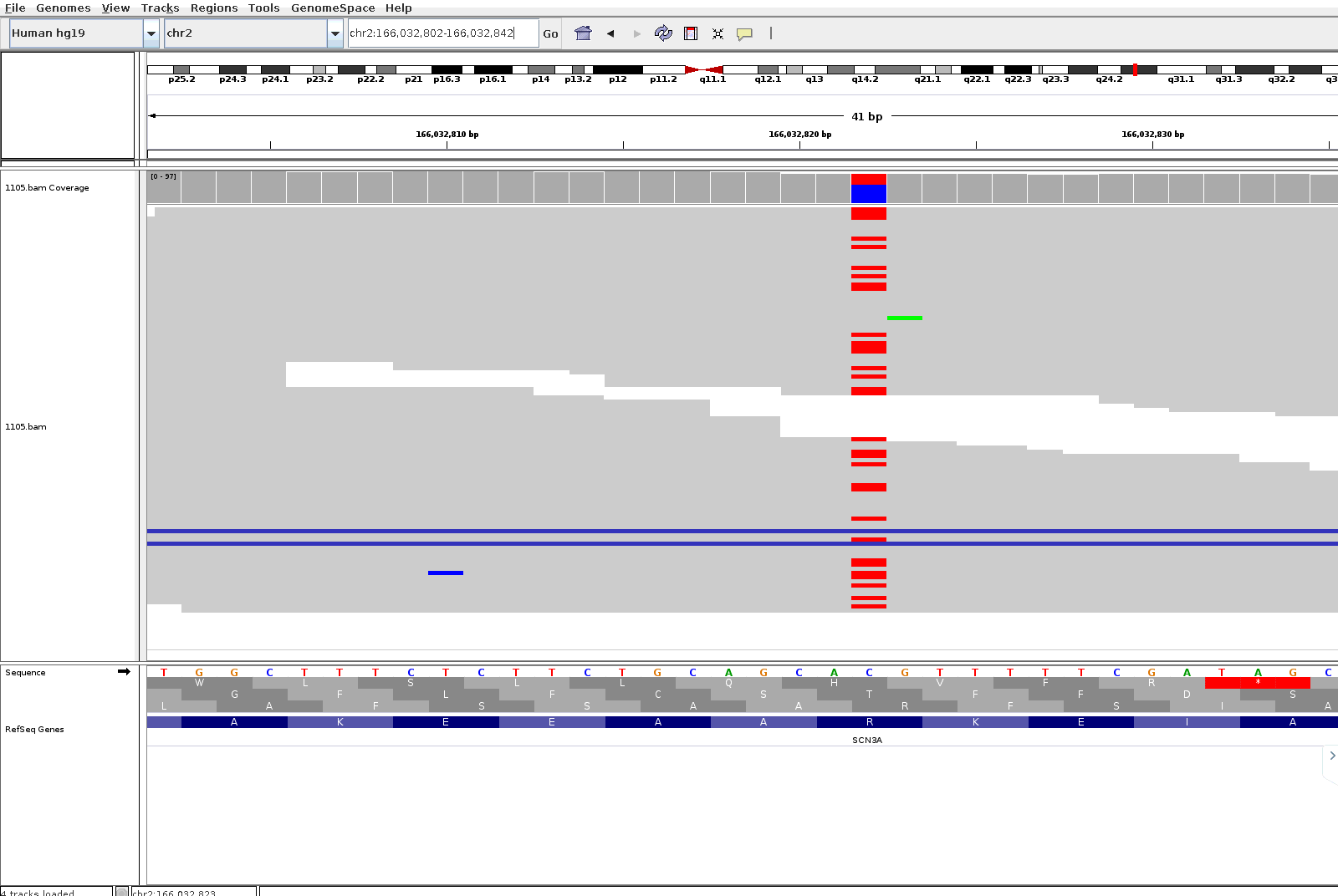

Figure 2F: The alternate allele T is present in gene SCN3A as a nonsynonymous SNV on chromosome 2 at position 166032822 in a sample belonging to the Family D006

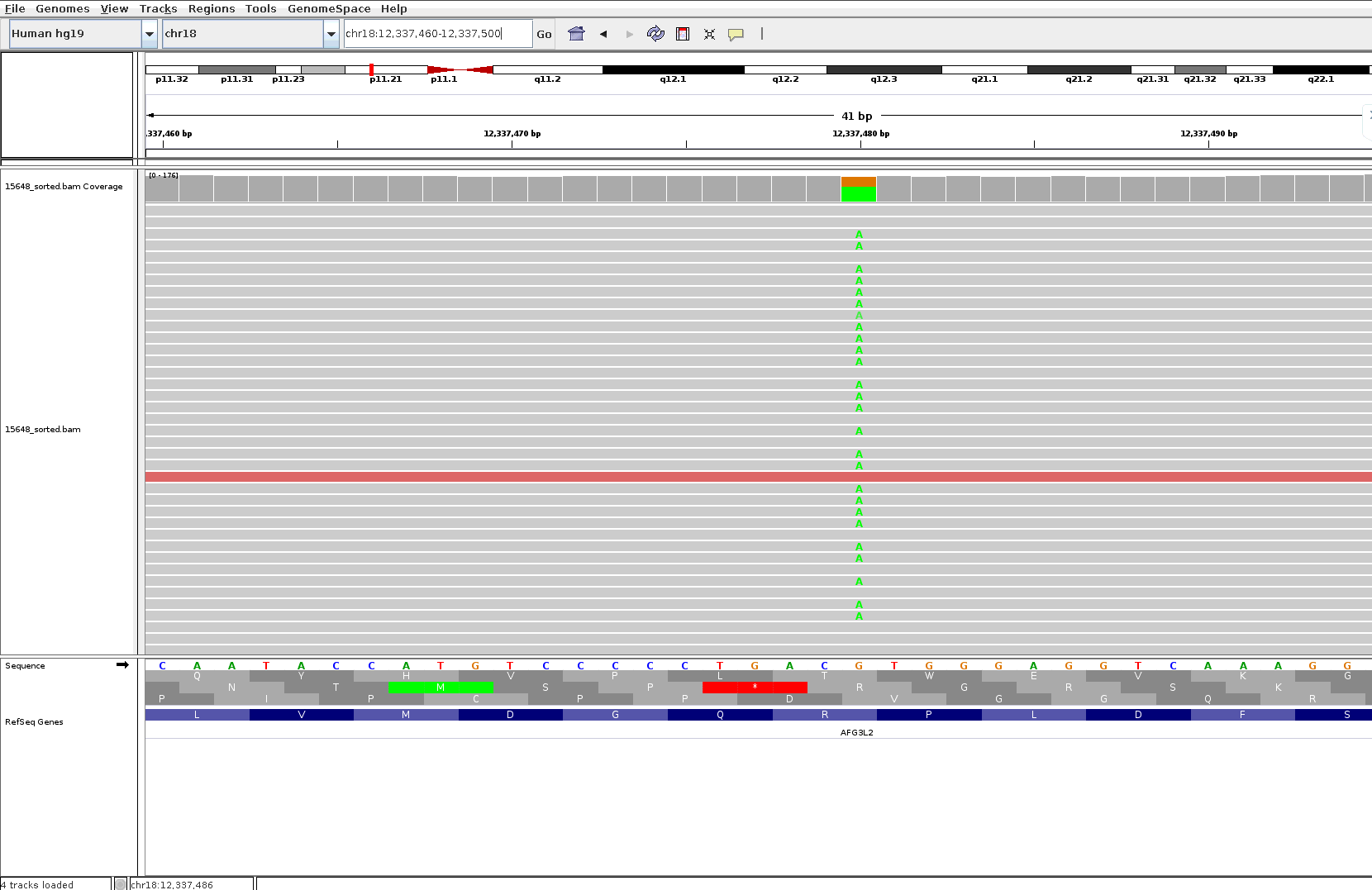

Figure 2G: The alternate allele A is present in gene AFG3L2 as a nonsynonymous SNV on chromosome 18 at position 12337480 in a sample belonging to the Family D002

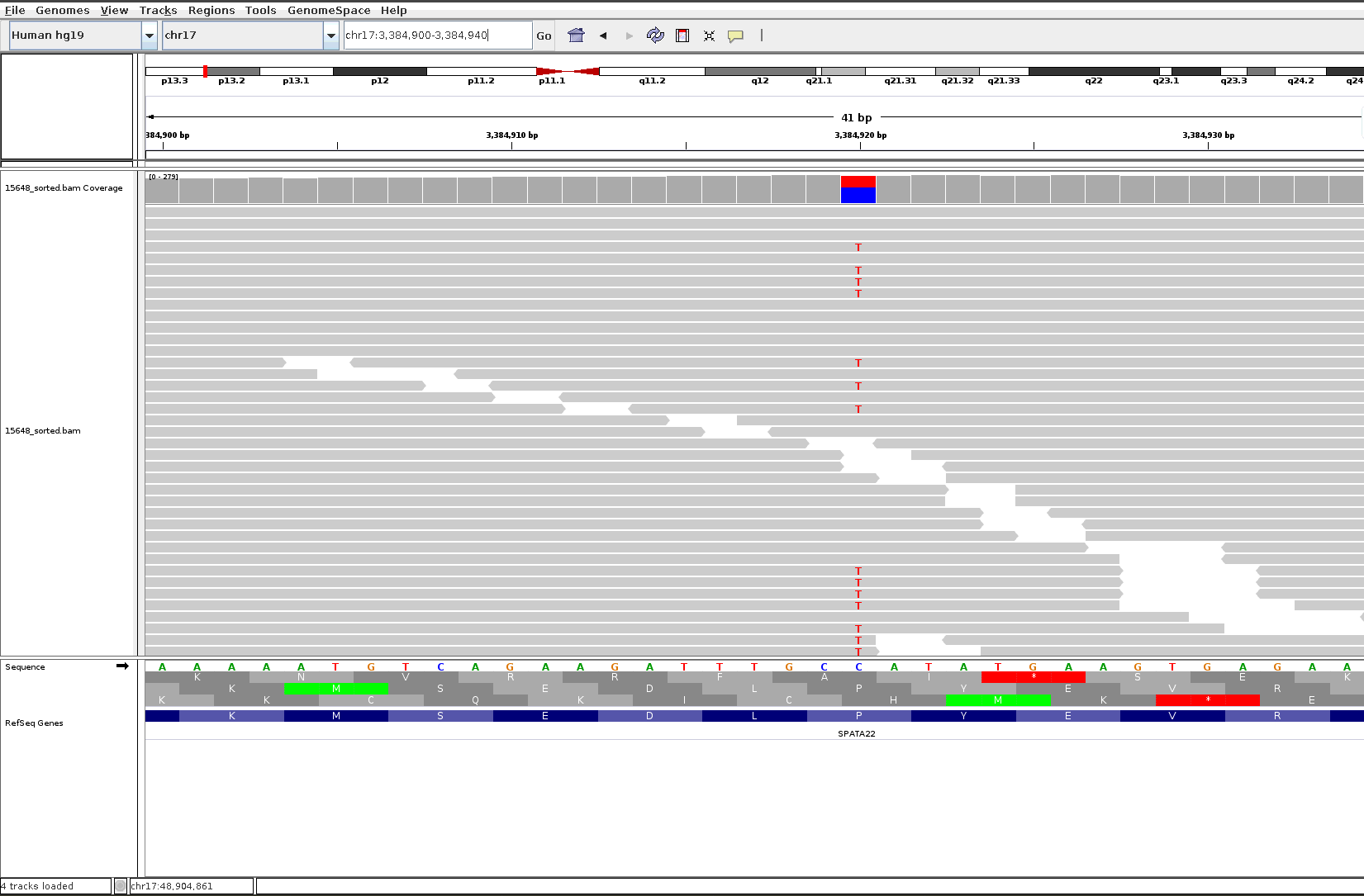

Figure 2H: The alternate allele T is present in gene ASPA as a nonsynonymous SNV on chromosome 17 at position 3384920 in a sample belonging to the Family D002

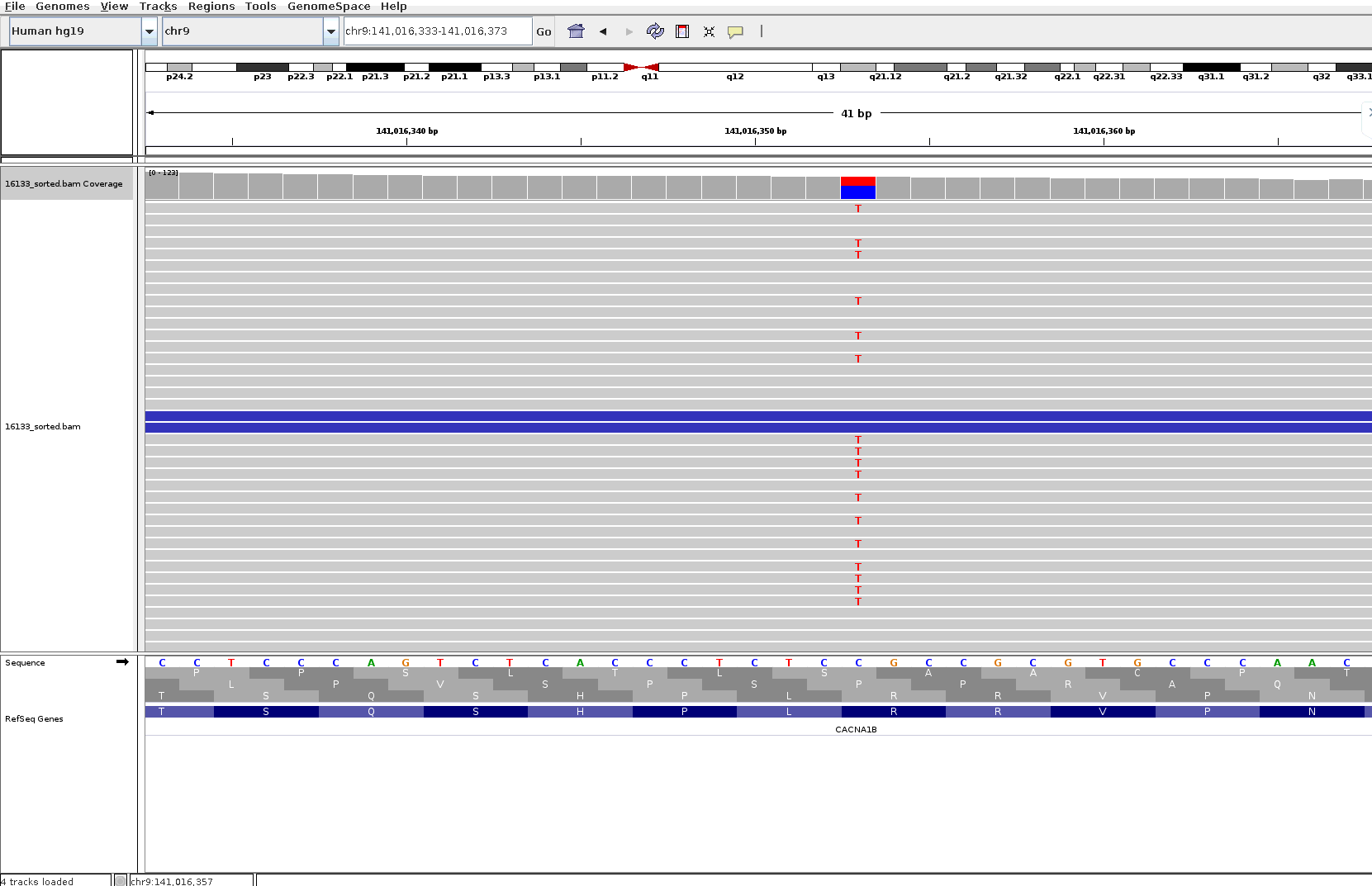

Figure 2i: The alternate allele T is present in gene CACNA1B as a nonsynonymous SNV on chromosome 9 at position 141016353 in a sample belonging to the Family D002

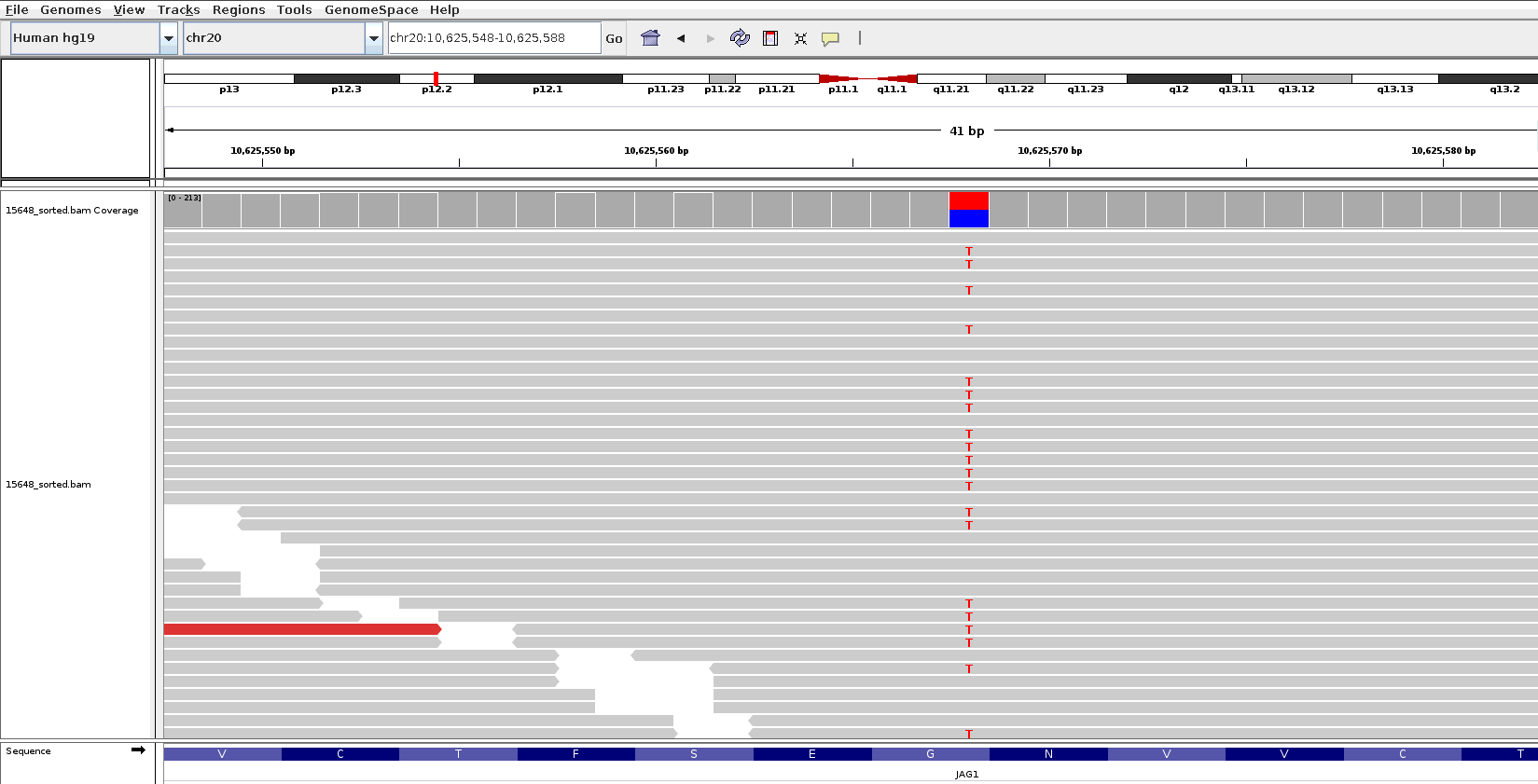

Figure 2j: The alternate allele T is present in gene JAG1 as a nonsynonymous SNV on chromosome 20 at position 10625568 in a sample belonging to the Family D002

**S5 Pedigrees included in the within pedigree prioritization**

Pedigrees D001 to D016 are attached as a supplementary document. The red circles indicate the cases and family controls sequenced in the present study. Each sequenced individual is numbered and the table details the diagnosis or the control status.

References:

1. Li M, Shen L, Chen L, Huai C, Huang H, Wu X, Yang C, Ma J, Zhou W, Du H, Fan L, He L, Wan C, Qin S. Novel genetic susceptibility loci identified by family based whole exome sequencing in Han Chinese schizophrenia patients. Transl Psychiatry. 2020;10:5.

2. Legge SE, Santoro ML, Periyasamy S, Okewole A, Arsalan A, Kowalec K. Genetic architecture of schizophrenia: a review of major advancements. Psychol Med. 2021:1-10.

3. Howrigan DP, Rose SA, Samocha KE, Fromer M, Cerrato F, Chen WJ, Churchhouse C, Chambert K, Chandler SD, Daly MJ, Dumont A, Genovese G, Hwu HG, Laird N, Kosmicki JA, Moran JL, Roe C, Singh T, Wang SH, Faraone SV, Glatt SJ, McCarroll SA, Tsuang M, Neale BM. Exome sequencing in schizophrenia-affected parent-offspring trios reveals risk conferred by protein-coding de novo mutations. Nat Neurosci. 2020;23:185-193.

4. Wilfert AB, Turner TN, Murali SC, Hsieh P, Sulovari A, Wang T, Coe BP, Guo H, Hoekzema K, Bakken TE, Winterkorn LH, Evani US, Byrska-Bishop M, Earl RK, Bernier RA, Consortium S, Zody MC, Eichler EE. Recent ultra-rare inherited variants implicate new autism candidate risk genes. Nat Genet. 2021;53:1125-1134.

5. Forstner AJ, Fischer SB, Schenk LM, Strohmaier J, Maaser-Hecker A, Reinbold CS, Sivalingam S, Hecker J, Streit F, Degenhardt F, Witt SH, Schumacher J, Thiele H, Nurnberg P, Guzman-Parra J, Orozco Diaz G, Auburger G, Albus M, Borrmann-Hassenbach M, Gonzalez MJ, Gil Flores S, Cabaleiro Fabeiro FJ, Del Rio Noriega F, Perez Perez F, Haro Gonzalez J, Rivas F, Mayoral F, Bauer M, Pfennig A, Reif A, Herms S, Hoffmann P, Pirooznia M, Goes FS, Rietschel M, Nothen MM, Cichon S. Whole-exome sequencing of 81 individuals from 27 multiply affected bipolar disorder families. Transl Psychiatry. 2020;10:57.

6. Toma C, Shaw AD, Allcock RJN, Heath A, Pierce KD, Mitchell PB, Schofield PR, Fullerton JM. An examination of multiple classes of rare variants in extended families with bipolar disorder. Transl Psychiatry. 2018;8:65.

7. Sul JH, Service SK, Huang AY, Ramensky V, Hwang SG, Teshiba TM, Park Y, Ori APS, Zhang Z, Mullins N, Olde Loohuis LM, Fears SC, Araya C, Araya X, Spesny M, Bejarano J, Ramirez M, Castrillon G, Gomez-Makhinson J, Lopez MC, Montoya G, Montoya CP, Aldana I, Escobar JI, Ospina-Duque J, Kremeyer B, Bedoya G, Ruiz-Linares A, Cantor RM, Molina J, Coppola G, Ophoff RA, Macaya G, Lopez-Jaramillo C, Reus V, Bearden CE, Sabatti C, Freimer NB. Contribution of common and rare variants to bipolar disorder susceptibility in extended pedigrees from population isolates. Transl Psychiatry. 2020;10:74.

8. Halvorsen M, Samuels J, Wang Y, Greenberg BD, Fyer AJ, McCracken JT, Geller DA, Knowles JA, Zoghbi AW, Pottinger TD, Grados MA, Riddle MA, Bienvenu OJ, Nestadt PS, Krasnow J, Goes FS, Maher B, Nestadt G, Goldstein DB. Exome sequencing in obsessive-compulsive disorder reveals a burden of rare damaging coding variants. Nat Neurosci. 2021;24:1071-1076.

9. Palmer DS, Howrigan DP, Chapman SB, Adolfsson R, Bass N, Blackwood D, Boks MPM, Chen C-Y, Churchhouse C, Corvin AP, Craddock N, Curtis D, Di Florio A, Dickerson F, Goes FS, Jia X, Jones I, Jones L, Jonsson L, Kahn RS, Landén M, Locke A, McIntosh A, McQuillin A, Morris DW, O’Donovan MC, Ophoff RA, Owen MJ, Pedersen N, Posthuma D, Reif A, Risch N, Schaefer C, Scott L, Singh T, Smoller JW, Solomonson M, St. Clair D, Stahl EA, Vreeker A, Walters J, Wang W, Watts NA, Yolken R, Zandi P, Neale BM. Exome sequencing in bipolar disorder reveals shared risk gene &lt;em&gt;AKAP11&lt;/em&gt; with schizophrenia. medRxiv. 2021:2021.2003.2009.21252930.

10. Jansen IE, Savage JE, Watanabe K, Bryois J, Williams DM, Steinberg S, Sealock J, Karlsson IK, Hagg S, Athanasiu L, Voyle N, Proitsi P, Witoelar A, Stringer S, Aarsland D, Almdahl IS, Andersen F, Bergh S, Bettella F, Bjornsson S, Braekhus A, Brathen G, de Leeuw C, Desikan RS, Djurovic S, Dumitrescu L, Fladby T, Hohman TJ, Jonsson PV, Kiddle SJ, Rongve A, Saltvedt I, Sando SB, Selbaek G, Shoai M, Skene NG, Snaedal J, Stordal E, Ulstein ID, Wang Y, White LR, Hardy J, Hjerling-Leffler J, Sullivan PF, van der Flier WM, Dobson R, Davis LK, Stefansson H, Stefansson K, Pedersen NL, Ripke S, Andreassen OA, Posthuma D. Genome-wide meta-analysis identifies new loci and functional pathways influencing Alzheimer's disease risk. Nat Genet. 2019;51:404-413.

11. Prokopenko D, Morgan SL, Mullin K, Hofmann O, Chapman B, Kirchner R, Alzheimer's Disease Neuroimaging I, Amberkar S, Wohlers I, Lange C, Hide W, Bertram L, Tanzi RE. Whole-genome sequencing reveals new Alzheimer's disease-associated rare variants in loci related to synaptic function and neuronal development. Alzheimers Dement. 2021.

12. Ganesh S, Ahmed PH, Nadella RK, More RP, Seshadri M, Viswanath B, Rao M, Jain S, Consortium A, Mukherjee O. Exome sequencing in families with severe mental illness identifies novel and rare variants in genes implicated in Mendelian neuropsychiatric syndromes. Psychiatry Clin Neurosci. 2019;73:11-19.

13. Gulsuner S, Stein DJ, Susser ES, Sibeko G, Pretorius A, Walsh T, Majara L, Mndini MM, Mqulwana SG, Ntola OA, Casadei S, Ngqengelele LL, Korchina V, van der Merwe C, Malan M, Fader KM, Feng M, Willoughby E, Muzny D, Baldinger A, Andrews HF, Gur RC, Gibbs RA, Zingela Z, Nagdee M, Ramesar RS, King MC, McClellan JM. Genetics of schizophrenia in the South African Xhosa. Science. 2020;367:569-573.

14. Singh T, Neale BM, Daly MJ, on behalf of the Schizophrenia Exome Meta-Analysis C. Exome sequencing identifies rare coding variants in 10 genes which confer substantial risk for schizophrenia. medRxiv. 2020:2020.2009.2018.20192815.

15. Jia X, Goes FS, Locke AE, Palmer D, Wang W, Cohen-Woods S, Genovese G, Jackson AU, Jiang C, Kvale M, Mullins N, Nguyen H, Pirooznia M, Rivera M, Ruderfer DM, Shen L, Thai K, Zawistowski M, Zhuang Y, Abecasis G, Akil H, Bergen S, Burmeister M, Chapman S, DelaBastide M, Jureus A, Kang HM, Kwok PY, Li JZ, Levy SE, Monson ET, Moran J, Sobell J, Watson S, Willour V, Zollner S, Adolfsson R, Blackwood D, Boehnke M, Breen G, Corvin A, Craddock N, DiFlorio A, Hultman CM, Landen M, Lewis C, McCarroll SA, Richard McCombie W, McGuffin P, McIntosh A, McQuillin A, Morris D, Myers RM, O'Donovan M, Ophoff R, Boks M, Kahn R, Ouwehand W, Owen M, Pato C, Pato M, Posthuma D, Potash JB, Reif A, Sklar P, Smoller J, Sullivan PF, Vincent J, Walters J, Neale B, Purcell S, Risch N, Schaefer C, Stahl EA, Zandi PP, Scott LJ. Investigating rare pathogenic/likely pathogenic exonic variation in bipolar disorder. Mol Psychiatry. 2021.

16. OMIM: Online Mendelian Inheritance in Man. Johns Hopkins University (Baltimore, MD), McKusick-Nathans Institute of Genetic Medicine; 2018.
